# Ethnicity-specific alterations of plasma and hepatic lipidomic profiles are related to high NAFLD rate and severity in Hispanic Americans, a pilot study

**DOI:** 10.1101/2021.04.08.21254907

**Authors:** Tagreed A. Mazi, Kamil Borkowski, John W. Newman, Oliver Fiehn, Christopher L. Bowlus, Souvik Sarkar, Karen Matsukuma, Mohamed R. Ali, Dorothy A. Kieffer, Yu-Jui Y. Wan, Kimber L. Stanhope, Peter J. Havel, Valentina Medici

**Author notes:** Corresponding author; Valentina Medici M.D., F.A.A.S.L.D., University of California, Davis Department of Internal Medicine, Division of Gastroenterology and Hepatology 4150 V Street, Suite 3500, Sacramento, CA 95817.

## Abstract

Nonalcoholic fatty liver disease (NAFLD) is a progressive condition that includes steatosis (NAFL) and nonalcoholic steatohepatitis (NASH). In the U.S., Hispanics (HIS) are afflicted with NAFLD at a higher rate and severity compared to other ethnicities. To date, the mechanisms underlying this disparity have not been elucidated. In this pilot study, we compared untargeted plasma metabolomic profiles for primary metabolism, complex lipids, choline and related compounds between a group of HIS (n =7) and White Caucasian (CAU, n =8) subjects with obesity and biopsy-characterized NAFL to ethnicity-matched lean healthy controls (n =14 HIS and 8 CAU). We also compared liver and plasma metabolomic profiles in a group of HIS and CAU subjects with obesity and NASH of comparable NAFLD Activity Scores, to BMI-matched NASH-free subjects in both ethnicities. Results highlight signs of metabolic dysregulation observed in HIS, independent of obesity, including higher plasma triglycerides, acylcarnitines, and free fatty acids. With NASH progression, there were ethnicity-related differences in the hepatic profile, including higher free fatty acids and lysophospholipids seen in HIS, suggesting lipotoxicity is involved in the progression of NASH. We also observed greater hepatic triglyceride content, higher plasma triglyceride concentrations and lower hepatic phospholipids with signs of impaired hepatic mitochondrial β-oxidation. These findings provide preliminary evidence indicating ethnicity-related variations that could potentially modulate the risk for progression of NALD to NASH.

**Highlights:**

- Hispanic subjects show signs of metabolic dysregulations independent of obesity.
- In nonalcoholic fatty liver, plasma metabolomic profiles are differentially altered between ethnicities.
- In nonalcoholic steatohepatitis, the hepatic lipidomic profiles are differentially altered between ethnicities.
- In Hispanics, nonalcoholic steatohepatitis shows higher triglycerides and lipotoxic lipids with signs of impaired hepatic mitochondrial β-oxidation.
- Our results indicate evidence of ethnicity-related variations in the pathogenesis of nonalcoholic fatty liver disease.

**Graphical Abstract:** 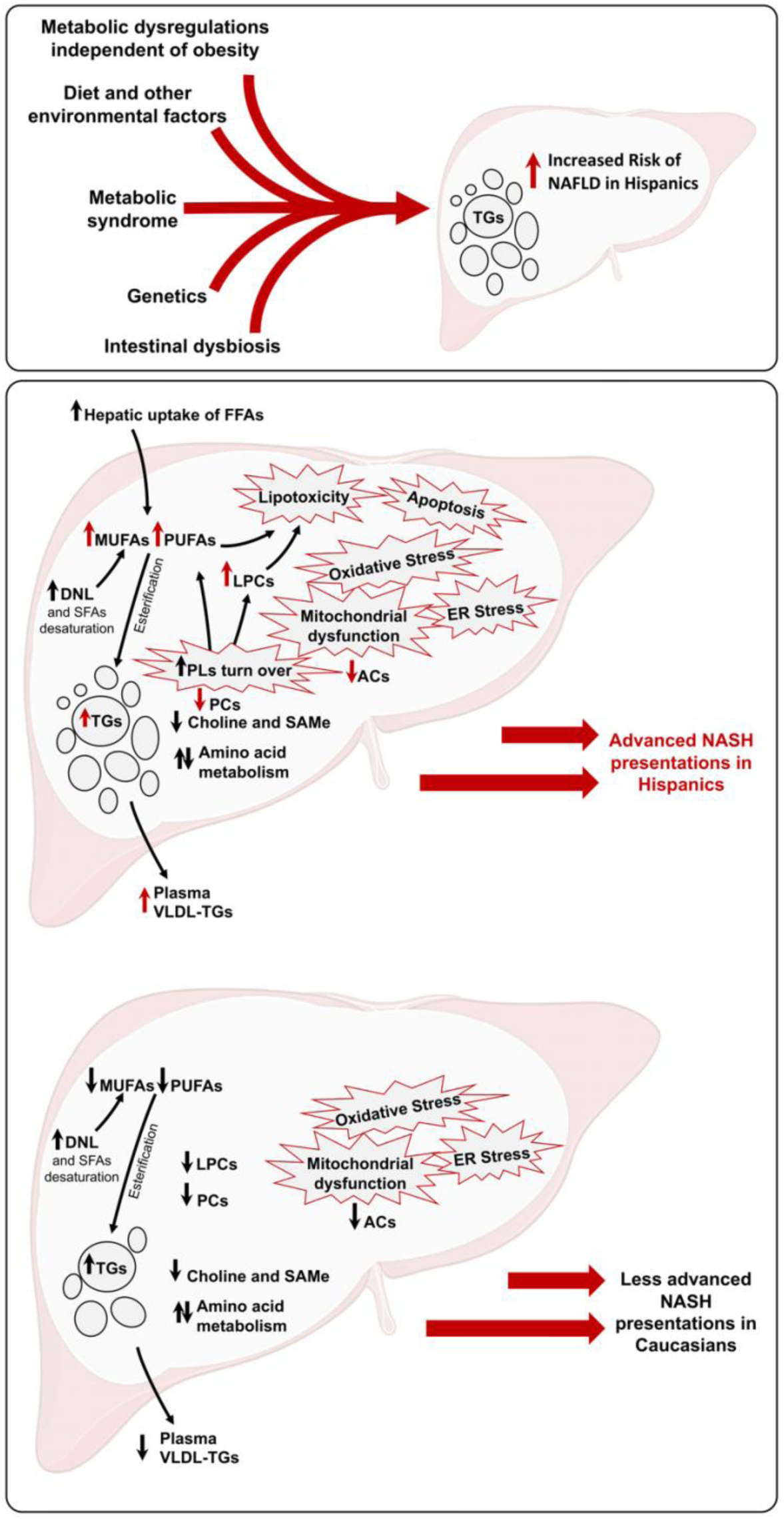

## Introduction

Nonalcoholic fatty liver disease (NAFLD) is a continuum of liver pathology that includes steatosis (NAFL), hepatocellular inflammation and ballooning, or nonalcoholic steatohepatitis (NASH) and cirrhosis [1]. It affects 25% of the general population and up to 95% of individuals with medically complicated obesity [2, 3]. Based on the strong correlations of NAFLD with metabolic syndrome and its components, NAFLD is argued to be its hepatic manifestation [4, 5]. In the U.S., disparity in NAFLD prevalence is reported, with Hispanics (HIS) being impacted at higher rate and severity compared to other ethnicities [6–11]. To date, the biological background underlying this disparity remains unclear.

NAFL develops when fatty acid availability, from the hepatic uptake of free fatty acids (FFA) and *de novo* lipogenesis (DNL), outweighs the utilization and disposal capacity, mainly by synthesis of triglycerides (TG) and other lipids, mitochondrial β-oxidation and export as very low-density lipoprotein [12]. Mechanisms of NAFLD progression are explained by the “multiple hit” model, in which lipid overload, the first hepatic hit, overwhelms the disposal, utilization and storage capacity. This results in a cascade of metabolic insults including oxidative stress, lipotoxicity, endoplasmic reticular stress, mitochondrial dysfunction and apoptosis, collectively, provoking hepatocyte injury, inflammation and disease progression [12, 13].

Mitochondrial function is altered in NAFLD and hepatic β-oxidation of fatty acids is reported to be both increased and decreased [14–16]. This inconsistency is thought to reflect a process of mitochondrial adaptation to compensate for the increased availability of FFAs, with an initial increase in β-oxidation, tricarboxylic acid cycle and oxidative phosphorylation [14, 15, 17]. Eventually, the metabolic capacity and antioxidant defenses are overwhelmed, resulting in hepatic oxidative stress and diverting FFAs to the production of hepatotoxic lipids [18, 19]. It is thought that accumulation and generation of hepatotoxic lipids is critical to NAFLD progression [20]. As mentioned above, lipotoxicity is considered a component of the “multiple hit” and may result from the accumulation of hepatotoxic lipids or intermediates, such as saturated fatty acids (SFA), ceramides (Cer) and lysophosphatidylcholine (LPC) [21–24]. Lipotoxicity can also result from a deficiency or imbalance in lipids essential for cellular integrity and function, including polyunsaturated fatty acids (PUFA) [24].

Previous metabolomic analysis of liver tissue and plasma have shown that NAFLD is associated with derangements of lipid, carbohydrate, and amino acid metabolism [25–29]. Dysregulation in one-carbon metabolism, including alterations in choline, betaine, methionine and in the universal methyl donor, S-Adenosyl-L-methionine (SAMe), is also implicated in NAFLD development and NASH severity, as shown from animal studies [30]. In humans, a recent metabolomic analysis identified a subtype of NAFLD subjects characterized by impaired one-carbon metabolism with NASH progression [31]. However, to our knowledge, no previous metabolomic profiling study has addressed ethnicity-related variations with regards to NAFLD.

The objective of this study was to investigate metabolomic profiles in a group of Hispanic (HIS) and White Caucasian (CAU) with obesity and biopsy-confirmed NAFLD. The identification of metabolomic differences between HIS and CAU subjects with NAFLD may provide a new research direction pointing towards ethnicity-specific changes as potential drivers for the disparity observed in NAFLD rate and progression. To this end, we employed untargeted metabolomic profiling of primary metabolism, complex lipids, choline and related metabolites in a group of Hispanic (NAFL-HIS) and Caucasian (NAFL-CAU) subjects with obesity and NAFLD of comparable histological presentations, as compared to a group of ethnicity-matched lean healthy control subjects. We also compared liver and plasma metabolomic profiles in a group of NASH (NASH-HIS) and (NASH-CAU) subjects with similar NAFLD Activity Score (NAS), to a group of BMI-matched NASH-free (0-NASH) subjects in both ethnicities.

## Subjects and methods

### 1. Subjects

Liver biopsies (n =17) and plasma samples (n =15) were retrieved from the biobank repository at the Division of Gastroenterology and Hepatology, UC Davis Medical Center. Samples were collected from bariatric surgery patients who self-identified ethnicity as either HIS or CAU. Included subjects were both males and females; age range 18−75 years; with class II and III obesity (body mass index (BMI) > 35.0 kg/m^2^); no previous diagnosis of acute or chronic diseases except for obesity or type 2 diabetes based on medical history. Exclusion criteria were diagnosis of secondary causes of chronic liver disease based on medical history, including viral and autoimmune hepatitis, HIV, hemochromatosis, alpha 1 anti-trypsin deficiency, Wilson disease and drug-induced liver disease; excessive alcohol intake (defined as >20 g/day for women and >30 g/day for men); and subjects with advanced fibrosis (stage >2). Healthy control subjects (HC; n =22) were recruited via public postings. Both male and female subjects were included; age between 18−75 years; BMI between 18 and 25 kg/m^2^; no current or previous diagnosis of chronic diseases; absence of signs of acute diseases.

For subjects with NAFLD, demographic, anthropometric and clinical data were collected retrospectively using data recorded within 7 days of bariatric surgery procedure. For HC subjects, demographic data were self-reported and anthropometric measurements were collected during one in-clinic visit. All subjects were consented with a signed form and the study protocol was approved by the Institutional Review Board at the University of California, Davis (protocol # 856052).

### 2. Biological samples

Liver tissue samples were collected by intraoperative core biopsy during bariatric surgery (time of collection is not available). For each subject, one sample was submitted in 10% formalin to the Department of Pathology for routine diagnostic interpretation. A second sample was flash-frozen in liquid nitrogen immediately after harvest and subsequently transferred and stored at −80 °C until analysis. Blood samples were collected by veno-puncture in EDTA-containing pre-cooled tubes after a 10-hour overnight fasting on the day of operation for NAFL group or in clinic for HC (time of collection is not available). Samples were centrifuged and stored at −80 °C until analysis.

### 3. Histopathology

Specimens received by the Department of Pathology were processed as per routine protocol. After diagnostic interpretation was completed, biopsy slides were retrieved and scored using the NASH Clinical Research Network (NASH-CRN) histology scoring system, which includes steatosis, hepatocellular ballooning, lobular inflammation, and fibrosis, by a gastrointestinal pathologist blinded to the subject’s clinical data. The NAFLD Activity Score (NAS) and fibrosis score were tabulated [32]. NASH diagnosis was determined by incorporating the histology scores into a diagnostic algorithm [33].

### 4. Untargeted metabolomics

Untargeted, semi-quantitative metabolomic profiling for plasma and liver samples was performed at the UC Davis West Coast Metabolomics Center. Samples were extracted using methanol: methyl tert-butyl ether (MTBE):water as previously described [34]. To profile metabolites related to primary metabolism including carbohydrates, sugar phosphates and amino acids, the aqueous phase was dried and subjected to trimethylsilation and methoximation and analyzed by gas chromatography/time-of-flight mass spectrometry (GC-TOF MS) [35]. Spectral data were processed and annotated using BinBase [36]. To profile FFAs and complex lipids, including mono-, di- and TGs, cholesteryl ester (CE), phospholipids (PL), sphingolipids (SL), the organic phase was dried, resuspended and analyzed by charged surface hybrid liquid chromatography/quadrupole time of flight mass spectrometry (CSH-QTOF MS/MS) [37]. Data collected in both positive and negative ion mode and processed using MassHunter (Agilent Technologies, Inc., Santa Clara, CA, http://www.agilent.com). Lipids were identified based on MS/MS fragmentation patterns using Lipidblast software [38]. To profile acylcarnitine, choline, betaine, S-Adenosyl-L-methionine (SAMe) and related metabolites, aqueous phase was analyzed by the biogenic amines platform using hydrophilic interaction liquid chromatography/quadrupole time of flight mass spectrometer (HILIC-QTOF MS/MS) [37]. Data were collected in both positive and negative mode. MS-DIAL was used for data processing [39]. For annotations, the three-levels of compound annotation by the Metabolomics Standards Initiative (MSI) was employed [40]. Metabolites with MSI level 1 annotation (i.e., an MS/MS spectral library with retention time and precursor mass) were excluded form statistical analyses. All mass spectra are available at Massbank of North America (http://mona.fiehnlab.ucdavis.edu). The study details are available on The Metabolomics Workbench (http://www.metabolomicsworkbench.org), ID number (ST000977).

In this study, fatty acids and complex lipids are described by name and lipid class, respectively, followed by number of (carbons, double bounds) of the fatty acyl moiety (i.e., linoleic acid (18:2n6) and TG(50-52:0-6)). The index of fatty acid desaturase-1(FADS1, Δ-5 desaturase) was estimated as the product to precursor ratio or arachidonic acid (20:4n6)/dihomo-γ-linolenic acid (20:3n6). Stearoyl-CoA desaturase (SCD1, or Δ-9 desaturase) was estimated as palmitoleic acid (16:1n7)/palmitic acid (16:0). The hepatic relative ratio of free n6 to n3 PUFAs was calculated as arachidonic acid (20:4n6)/eicosapentaenoic acid (20:5n3) and arachidonic acid (20:4n6)/docosahexaenoic acid (22:6n3).

### 5. Statistical analysis

Statistical analysis was performed using JMP Pro 14.1 (SAS Institute Inc., Cary, NC; http://www.jmp.com). Unknowns and metabolites and with MSI level 1 identification from HILIC-QTOF MS/MS platform were excluded. Outliers were detected and excluded using the robust Huber M test and missing data were imputed using multivariate normal imputation if greater than 70% complete.

Metabolites with more than 30% missing data were excluded. The Johnson’s transformation was used to attain normal data distributions for statistical assessment. Metabolite means were calculated using Log normalized data, fold changes were calculated as the mean of NAFL/HC for comparison between NAFL and HC or NASH/0-NASH for comparison between NASH and 0-NASH. Students’ *t*-test was used to compare means. Differences were considered likely at *p* <0.05, and possible if the *p*-value was ≥0.05 and <0.1. Full factorial analysis of covariates (ANCOVA) was employed to evaluate the interaction of ethnicity x health status (i.e., NASH or NAFL). This model included ethnicity, health status, ethnicity x health status interaction as fixed effects, and age and sex as covariates. Benjamini-Hochberg false discovery rate (FDR) correction for multiple comparisons using a *q* =0.2 was performed, given the pilot nature and small sample number of this study [41]. To reduce data dimensionality and facilitate visualization, metabolites were clustered using the JMP implementation of the SAS VARCLUS procedure, a principal component analysis (PCA)-based clustering algorithm, with cluster components calculated as the linear sum of all variables in a cluster. For NAFL vs. HC comparisons, metabolites with *p* <0.1 for ethnicity x NAFL interaction were clustered. For NASH vs. 0-NASH comparisons, metabolites with *p* <0.05 for the NASH effect within ethnicity groups and with *p* <0.05 for the ethnicity x NASH interaction were clustered.

To focus on the differential effect of NAFL between ethnicities, cluster components were subjected to partial least square-discriminant analysis (PLS-DA) separately in each ethnicity with leave-one-out cross validation (LOOCV) [42]. A variable importance in projection (VIP) score of >1 was set as a threshold for variable selection [43]. Because the PLS-DA model was constructed using reduced set of data to highlight only ethnicity-specific differences between NAFL and HC, the VIP scores should not be directly compared between the two analyses.

To further examine the effect of ethnicity between NASH and 0-NASH, cluster components were reevaluated using *t*-test for group comparison within ethnicity and ANCOVA for ethnicity x NASH. To examine the association between metabolomic profiles and liver histological scoring, Spearman’s rank correlations was performed. To correct for multiple testing, Benjamini-Hochberg FDR adjustment was performed with *q* =0.2 [41].

### 6. Over representation analysis

To provide an overview on the general changes in metabolomic profile, we employed ChemRICH [44], a statistical enrichment analysis based on chemical similarity. This approach clusters metabolites into non-overlapping chemical groups using Tanimoto substructure chemical similarity coefficients and calculates cluster *p*-values using the Kolmogorov–Smirnov test.

### 7. Pathway enrichment analysis

Pathway analysis was performed using MetaboAnalyst (McGill University, Quebec, CA; http://metaboanalyst.ca) [45]. Metabolites with differential alterations between ethnicity in NASH (*p* <0.05) were compared against pathway-associated metabolite sets from Kyoto Encyclopedia of Genes and Genomes (KEGG) [46]. Fisher’s Exact test was used to assess over-representation and the relative betweenness centrality was used for topology analysis.

## Results

### 1. Subject characteristics

The clinical and histological features for NAFL and NASH subjects are presented in Table 1 and Table 2. When comparing between ethnicities, both NAFL and NASH groups showed no difference in age, BMI and other clinical parameters including histology NAS scores.

**Table 1.** Demographic, clinical and histological characteristics of study subjects. General characteristics are shown as percent (for categorical data) and mean ± SEM (for nominal data). Group comparisons were performed by chi-square test (categorical) or *t*-test (nominal). (a) NAFL-HIS vs. HC-HIS; (b) NAFL-CAU vs. HC-CAU; (c) NAFL-HIS vs. NAFL-CAU. ALT, alanine aminotransferase; AST, aspartate aminotransferase; BMI, body mass index; DM, diabetes mellitus; FBG, fasting blood glucose; HbA1c, hemoglobin A1c; HDL, high-density lipoprotein; LDL, low density lipoprotein; NAS, the NAFLD Activity Score; TG, triglycerides.

|  | Hispanics |  | Caucasians |  | *p*-value |  |  |
| --- | --- | --- | --- | --- | --- | --- | --- |
|  | NAFL | HC | NAFL | HC | *a* | *b* | *c* |
| Liver, *n* (F/M) | 7 (5/2) | - | 10 (5/5) | - |  |  |  |
| Plasma, *n* (F/M) | 7 (5/2) | 14 (6/8) | 8 (4/4) | 8 (4/4) |  |  |  |
| Age (Yrs.) | 45 ± 16 | 43 ± 14 | 50 ± 16 | 44 ± 12 | 0.818 | 0.413 | 0.55 |
| BMI (Kg/m<sup>2</sup>) | 47 ± 7 | 26 ± 2 | 42 ± 7 | 25 ± 3 | <.0001 | <.0001 | 0.28 |
| DM, yes (%) | 3 (42.9) | - | 4 (40) | - | - | - | 0.913 |
| FBG mmol/L | 93 ± 5 | - | 94 ± 13 | - | - | - | 0.88 |
| Cholesterol (mg/dL) | 168 ± 29 | - | 173 ± 33 | - | - | - | 0.952 |
| TG (mg/dL) | 112 ± 37 | - | 139 ± 65 | - | - | - | 0.42 |
| HDL (mg/dL) | 44 ± 7 | - | 41 ± 8 | - | - | - | 0.403 |
| LDL (mg/dL) | 102 ± 23 | - | 104 ± 33 | - | - | - | 0.855 |
| HbA<sub>1c</sub> (%) | 6.1 ± 0.7 | - | 6.2 ± 0.8 | - | - | - | 0.67 |
| AST (U/L) | 27 ± 12 | - | 31 ± 19 | - | - | - | 0.499 |
| ALT (U/L) | 34 ± 18 | - | 31 ± 10 | - | - | - | 0.752 |
| Platelet | 308 ± 58 | - | 292 ± 100 | - | - | - | 0.506 |
| NAS | 2.43 ± 2.40 | - | 3 ± 1.5 | - | - | - | 0.653 |
| NASH, yes (%) | 6 (60) | - | 5 (50) | - | - | - | 0.708 |
| Steatosis (%) |  |  |  |  |  |  |  |
| < 5% | 3 (42.8) | - | 2 (20) | - | - | - | 0.452 |
| 5 to ≤ 33% | 2 (28.6) | - | 5 (50) | - | - | - |  |
| 34 to ≤ 66 % | 2 (28.6) | - | 2 (20) | - | - | - |  |
| > 66% | 0 (0) | - | 1 (10) | - | - | - |  |
| Inflammation (%) |  |  |  |  |  |  |  |
| None | 3 (42.9) | - | 1 (10) | - | - | - | 0.763 |
| Mild | 1 (14.3) | - | 6 (60) | - | - | - |  |
| Moderate | 2 (28.6) | - | 3 (30) | - | - | - |  |
| Severe | 1 (14.3) | - | 0 (0) | - | - | - |  |
| Ballooning (%) |  |  |  |  |  |  |  |
| None | 4 (57) | - | 5 (50) | - | - | - | 0.772 |
| Few | 3 (42.9) | - | 5 (50) | - | - | - |  |
| Many | 0 (0) | - | 0 (0) | - | - | - |  |
| Fibrosis (%) |  |  |  |  |  |  |  |
| None | 7 (100) | - | 9 (90) | - | - | - | 0.998 |
| 1A | 0 (0) | - | 1 (10) | - | - | - |  |
| 2 | 0 (0) | - | 0 (0) | - | - | - |  |
| 4 | 0 (0) | - | 0 (0) | - | - | - |  |

**Table 2.** Demographic, clinical and histological characteristics of subjects with NASH. General characteristics are shown as percent (for categorical data) and mean ± SEM (for nominal data). Comparison were performed by *t*-test (nominal) or chi-square test (categorical). (a) NASH-HIS vs. 0-NASH-HIS; (b) NASH-CAU vs. 0-NASH-CAU. ALT, alanine aminotransferase; AST, aspartate aminotransferase; BMI, body mass index; FBG, fasting blood glucose; NAS, the NAFLD Activity Score; NASH, Nonalcoholic steatohepatitis; 0-NASH, Nonalcoholic steatohepatitis-free; TG, triglycerides.

|  | Hispanics |  | Caucasians |  | *p*-value |  |
| --- | --- | --- | --- | --- | --- | --- |
|  | 0 NASH | NASH | 0 NASH | NASH | *a* | *b* |
| Liver, *n* (F/M) | 4(3/1) | 3(2/1) | 5(3/2) | 5(2/3) |  |  |
| Plasma, *n* (F/M) | 4(3/1) | 3(2/1) | 5(3/2) | 3(1/2) |  |  |
| Age (Yrs.) | 45 ± 21 | 46 ± 10 | 54 ± 19 | 45 ± 14 | 0.872 | 0.405 |
| BMI (Kg/m<sup>2</sup>) | 47.4 ± 7.9 | 45.7 ± 6.7 | 39.9 ± 4.4 | 44.4 ± 3.8 | 0.787 | 0.406 |
| FBG mmol/L | 93 ± 5 | 94 ± 4 | 98 ± 15 | 90 ± 10 | 0.873 | 0.379 |
| TG (mg/dL) | 105 ± 39 | 121 ± 40 | 120 ± 80 | 157 ± 50 | 0.59 | 0.35 |
| AST (U/L) | 20 ± 3 | 36 ± 14 | 21 ± 3 | 40 ± 23 | 0.064 | **0.0276** |
| ALT (U/L) | 23 ± 9 | 48 ± 18 | 25 ± 2.2 | 38 ± 11 | **0.0431** | **0.0246** |
| Steatosis | 12 ± 19 | 22.7 ± 9.3 | 15.6 ± 15.5 | 32.2 ± 21.8 | 0.197 | 0.1474 |
| NAS | 0.75 ± 1.1 | 4.7 ± 0.6 | 1.8 ± 1 | 4 ± 1 | **0.004** | **0.017** |
| Inflammation (%) |  |  |  |  |  |  |
| None | 3(75) | 0(0) | 1(20) | 0(0) |  |  |
| Mild | 1(25) | 0(0) | 3(60) | 3(60) |  |  |
| Moderate | 0(0) | 2(66.7) | 1(20) | 2(40) | **0.998** | 0.323 |
| Severe | 0(0) | 1(33.3) | 0(0) | 0(0) |  |  |
| Ballooning (%) |  |  |  |  |  |  |
| None | 4(100) | 0(0) | 5(100) | 0(0) |  |  |
| Few | 0(0) | 3(100) | 0(0) | 5(100) | **<0.0001** | **<0.0001** |
| Many | 0(0) | 0(0) | 0(0) | 0(0) |  |  |
| Fibrosis (%) |  |  |  |  |  |  |
| None | 4(100) | 3(100) | 5(100) | 4(80) |  |  |
| 1A | 0(0) | 0(0) | 0(0) | 1(20) |  |  |
| 2 | 0(0) | 0(0) | 0(0) | 0(0) | 1 | **0.998** |
| 3 | 0(0) | 0(0) | 0(0) | 0(0) |  |  |
| 4 | 0(0) | 0(0) | 0(0) | 0(0) |  |  |

In NAFL groups, the mean age was 45 ± 16 and 50 ± 16 years in HIS and CAU, respectively (*p* >0.05); 29% of HIS and 30% of CAU had moderate to severe steatosis; 43% of HIS and 50% of CAU were diagnosed with NASH and exhibited various degrees of lobular inflammation. One NAFL-CAU subject had portal/periportal fibrosis, or stage 1A. The mean NAS score for NAFL subjects was 2.4 ± 2.4 and 2.9 ± 1.5 for HIS and CAU, respectively (*p* >0.05). When compared to HC, BMI was 80% higher in NAFL-HIS and 70% higher in NAFL-CAU. In NASH subjects, the mean NAS score was 4.7 ± 0.58 and 4 ± 1 for HIS and CAU, respectively (*p* >0.05). Compared to 0-NASH, both ethnicities showed no difference in age, BMI or other clinical parameters.

### 2. Ethnicity-specific differences in plasma metabolome in NAFL

Given the low sample size and broad analysis in this study, we first evaluated NAFL-dependent changes by chemical classes separately in each ethnicity, effectively reducing the number of comparisons (Fig. 1). In both ethnicities, when compared to corresponding HC, NAFL was associated with lower plasma phosphatidylcholines (PC), unsaturated LPCs, galactosyl Cers, CEs and amino acids. Specific to CAU, NAFL showed higher levels of ACs and unsaturated FFAs, driven by elevated PUFAs including linoleic acid (18:2n6), α-linolenic acid (18:3n3), eicosadienoic acid (20:2n6) and MUFAs, heptadecenoic acid (17:1n7) and physeteric acid (14:1n7). Also, NAFL-CAU showed a reduction in many PLs including ether-linked PLs (Table S1). These differences in ACs and in unsaturated fatty acids were not evident in the NAFL-HIS group, which was characterized by alterations in sphingomyelins (SM) and xanthines, with higher level hydroxybutyrates, trimethyl ammonium compounds and sugar alcohols (Table S2).

**Fig 1.**
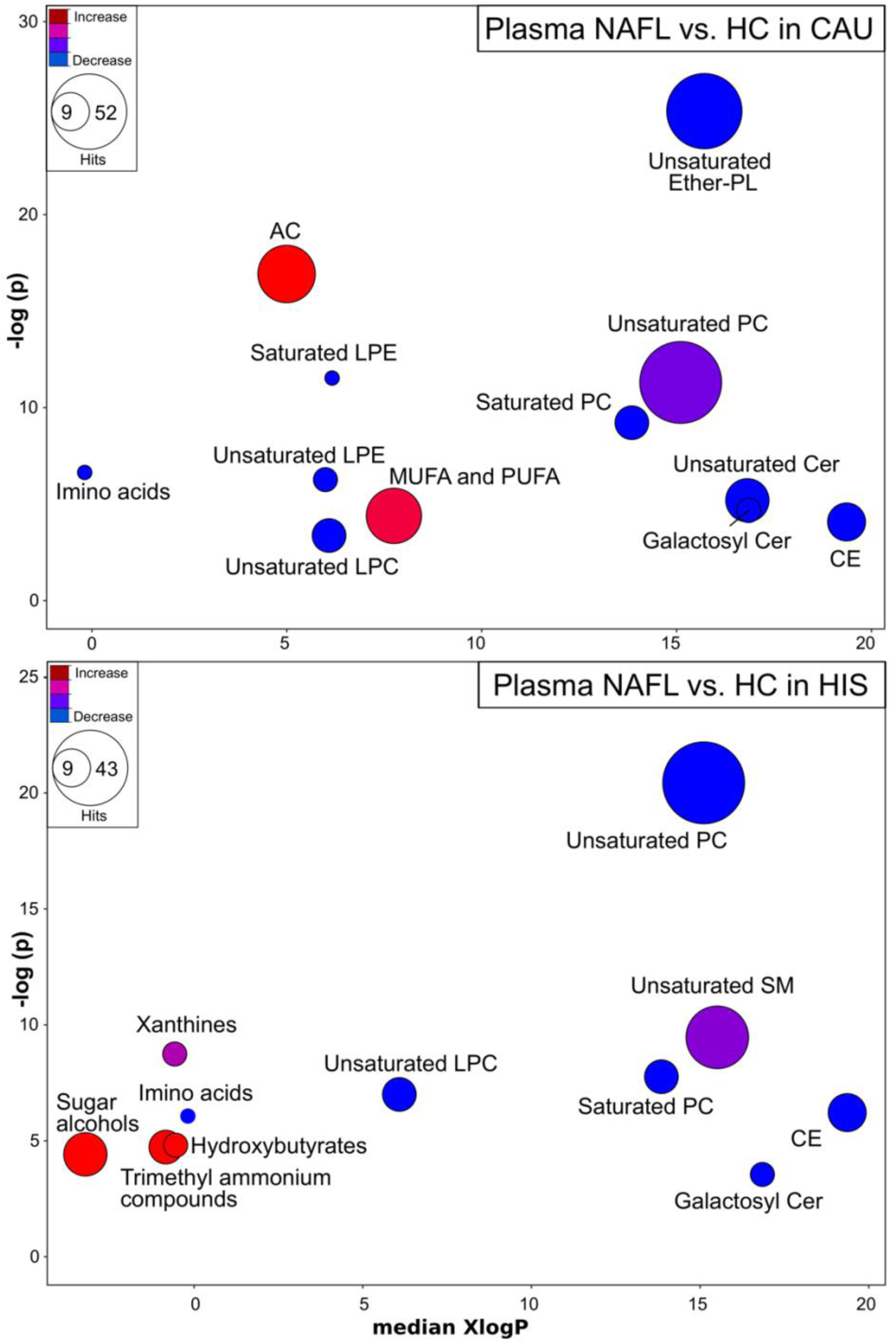
Plasma metabolites altered by chemical class in NAFL, compared to HC in both ethnicities. Chemical similarity enrichment analysis (ChemRICH) and enrichment statistics plot for NAFL vs. HC in CAU (top panel) and HIS (bottom panel). Each cluster represents altered chemical class of metabolites (*p* <0.05). Cluster sizes represent the total number of metabolites. Cluster’s color represents the directionality of metabolite differences: red – higher in NAFL; blue – lower in NAFL. Colors in between refer to mixed population of metabolites manifesting both higher and lower levels in NAFL when compared to the control. The *x*-axis represents the cluster order on the chemical similarity tree. The plot *y*-axis shows chemical enrichment *p*-values calculated using Kolmogorov–Smirnov test. Only clusters with *p* <0.05 are shown. FDR-adjustment *q* =0.2, and clusters with FDR-adjusted *p* ≥0.2 are shown in gray. (*n*, NAFL-HIS =7, HC-HIS =14; NAFL-CAU =8, HC-CAU =8). The detailed ChemRICH results are shown in (Table S1 and S2). AC, Acylcarnitines; CAU, White Caucasian; Cer, Ceramides; CE, Cholesteryl ester; DG, Diglycerides; Ether-PL, Ether-linked phospholipids; HIS, Hispanic; LPC, Lysophosphatidylcholine; LPE, Lysophosphatidylethanolamines; MUFA, Monounsaturated fatty acid; PC, Phosphatidylcholines; PUFA, Polyunsaturated fatty acids; SFA, Saturated fatty acids; SM, Sphingomyelins; NAFL, Steatosis; TG, Triglycerides.

To further examine this ethnicity-associated divergence in metabolomic profile, ANCOVA was performed with interaction (ethnicity x NASH). Of the 940 metabolites detected, 46 plasma metabolites (4.9%) were found altered (*p* <0.05) in NAFL between ethnicities with an additional 59 metabolites showing a *p*-value between 0.05 and <0.1. However, none of these passed the FDR-based multiple comparisons adjustment. Although the probability of differences was low and the number of findings was close to that expected by random chance, due to the strong class-specific changes and the exploratory nature of this study, we proceeded with a characterization of metabolite differences between the groups.

The variable clustering approach reduced the data to 21 cluster components. To highlight the apparent ethnicity-specific metabolic differences between NAFL and HC groups, these cluster components were projected using a PLS-DA (Fig. 2). In CAU, 11 of the 21 clusters contributed to the discrimination of NAFL and HC in CAU with VIP scores >1. These clusters were represented by ether-linked PL(32-40:0-5) (cluster 8 and 15), PE(18-36:0-1) and PC(34-36:0-3) (cluster 3) which were lower in NAFL-CAU, and PC(38:4-5) (cluster 20) which appeared higher. Also, NAFL-CAU showed higher abundance of short to medium length acylcarnitines (i.e. AC(2-14:0-2); (cluster 2 and 7) and in non-esterified FFAs (cluster 2), including the SFAs myristic acid (14:0) and margaric acid (17:0), oleic acid (18:1n9); the PUFAs linoleic acid (18:2n6) and α-linolenic acid (18:3n3). Similar trends were observed for docosahexaenoic acid (22:6n3) and eicosapentaenoic acid (20:5n3) (cluster 21), but with VIP <1. Also, NAFL-CAU showed higher levels of the intestinal microbiota-related, trimethylamine N-oxide (TMAO) and indole-3-acetate (cluster 5) and lower levels of the purine-related metabolites, adenosine and guanine (cluster 16 and 13).

**Fig 2.**
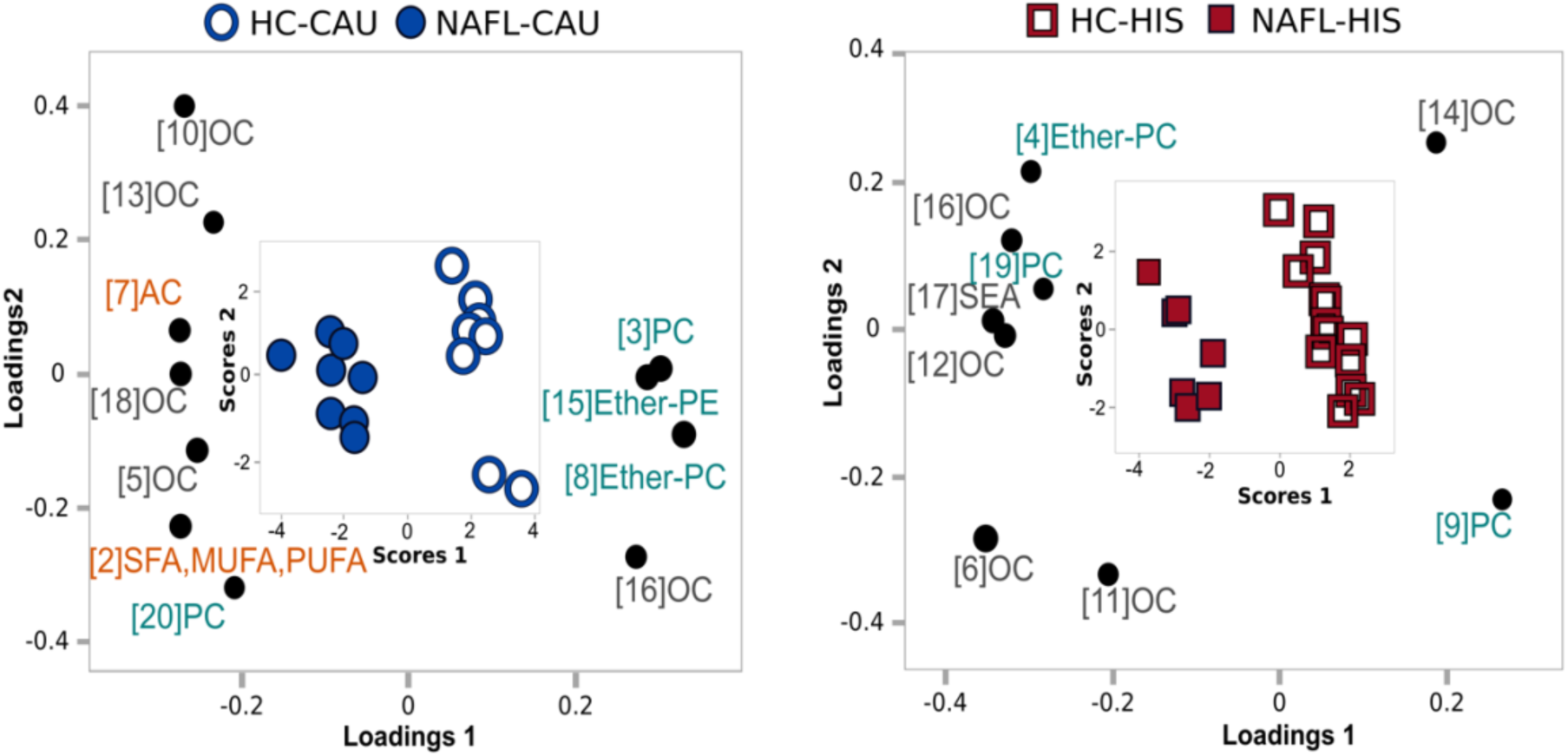
Supervised clustering model illustrating potential ethnicity-specific variations in plasma observed between NAFL vs. HC in both ethnicities. Metabolite clusters with potential ethnicity interaction (*p* <0.1) from ANCOVA were projected into partial least square-discriminant analysis (PLS-DA) model to highlight the nature and contribution of different cluster components into the separation between NAFL and HC in both ethnicities. Only clusters of variable importance in projection (VIP)>1 are illustrated. A combined loading and score plot for **a)** NAFL-CAU vs. HC-CAU; **b)** NAFL-HIS vs. HC-HIS. The model was validated with leave-one-out cross validation (LOOCV). The (Q^2^), (R^2^X) and (R^2^Y) is 0.817, 0.285 and 0.918; 0.816, 0.204 and 0.911 for CAU and HIS, respectively. The details on metabolites and clusters components are shown in (Table S3). AC, Acylcarnitines; CAU, White Caucasian; Ether-PC, Ether-linked phosphatidylcholines; Ether-PE, Ether-linked phosphatidylethanolamines; HC, Healthy control; HIS, Hispanic; MUFA, Monounsaturated fatty acid; OC, Organic compounds; PC, Phosphatidylcholines; PUFA, Polyunsaturated fatty acids; NAFL, Steatosis; SEA, Stearoyl ethanolamine; SFA, Saturated fatty acids.

The differences in ACs and in non-esterified FFAs were not evident in NAFL-HIS, as compared to corresponding HC. Instead, 9 of the 21 clusters contributed to the discrimination of NAFL and HC with VIP scores >1. These clusters indicated lower levels of PC(40:6-7) (cluster 9) and higher levels of PC(38:4) (cluster 19) and ether-linked PL (36:1-4) (cluster 4 and 19). There was also evidence of higher abundance of metabolites related to purine metabolism, xanthine and hypoxanthine (cluster 11) and in the endocannabinoid-like stearoylethanolamide (cluster 17), with differences in multiple other organic compounds (cluster 6, 11, 12, 14, 16). Because the PLS-DA was generated using a reduced number of metabolites (with ethnicity interaction *p* <0.1), the VIP scores should not be compared directly between the two models. Details on ANCOVA, variable clustering and PLS-DA results are shown in Table S3.

Together, results suggest metabolomic variation between NAFL and HC in CAU with alterations in PLs, ACs, non-esterified FFAs and some organic compounds, while in NAFL-HIS, changes were limited to PLs and some organic compounds. In addition, plasma FFA and AC profiles appear differentially altered with NAFL between ethnicity groups. These findings are depicted by the unsupervised PCA performed on cluster components (Fig. S1), showing complete separation of NAFL-CAU from HC-CAU, indicating these two groups are metabolically distinct, while there was a partial overlap between NAFL-HIS and HC-HIS, suggesting less metabolic variations between these groups.

### 3. High acylcarnitines, triglycerides and unsaturated fatty acids distinguish HIS from CAU in HC

The limited differences seen between NAFL and HC in HIS may reflect metabolic dysregulations in lean healthy HIS. To examine this, we compared plasma metabolomic profile in HC between ethnicities. Chemical similarity analysis clearly showed that in lean healthy subjects, HIS presented with higher ACs, TGs and unsaturated FFAs (Fig 3 and Table S4). HC-HIS had higher level (*p* <0.05) of the FFAs, margaric acid (17:0), oleic acid (18:1n9), linoleic acid (18:2n6), with similar trends in a number of other fatty acids. In addition, HC-HIS presented higher TG(51-58:1-9), DG(36-38:2-6), AC(2-16:0-1), altered PLs profiles, lower levels of the amino acids, glycine and alanine and the purine-related metabolites, adenosine, and hypoxanthine (Table S5).

**Fig 3.**
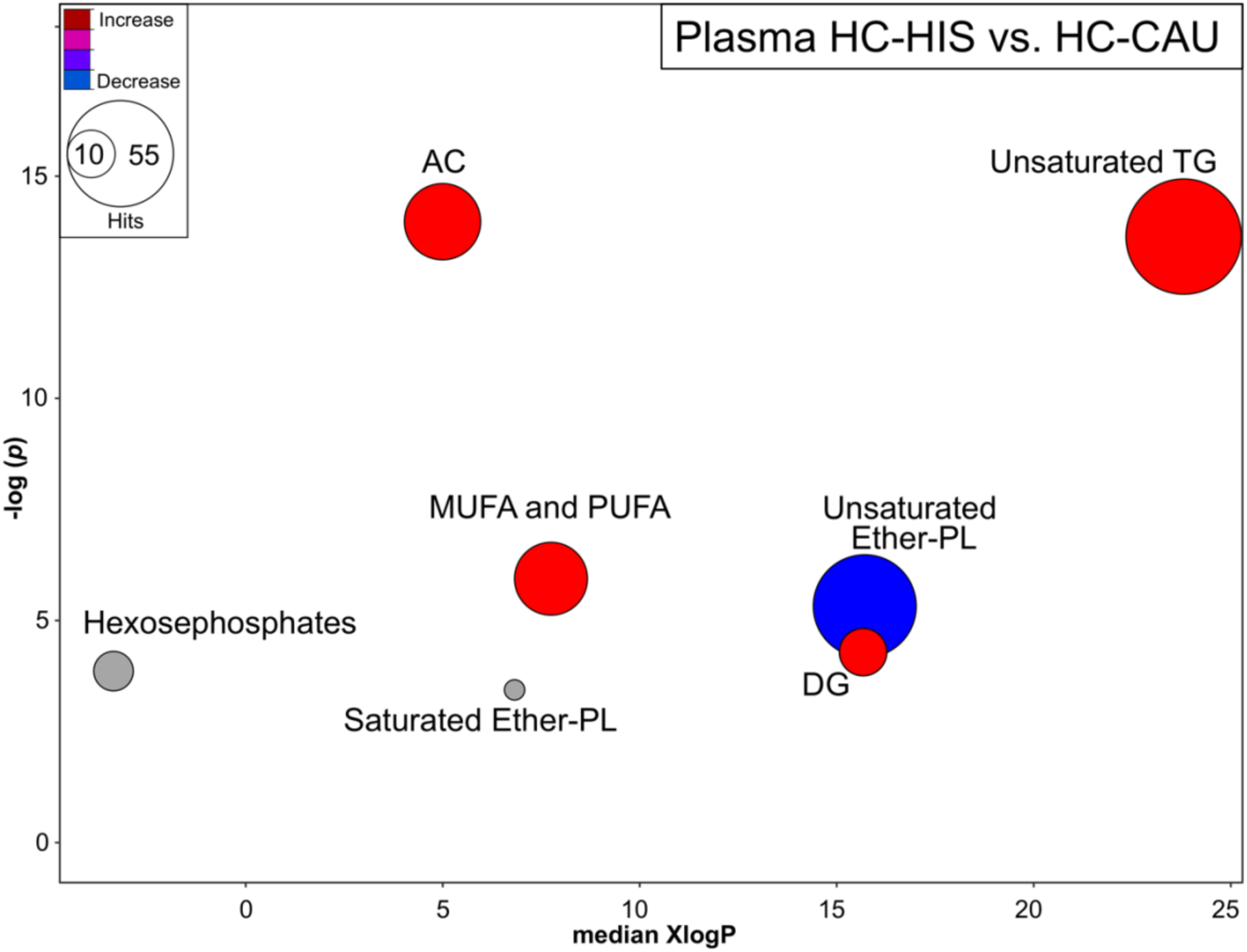
Plasma metabolites altered by chemical class in lean healthy control subjects between ethnicities. Chemical similarity enrichment analysis (ChemRICH) highlighting plasma metabolomic changes observed in HC-HIS, compared to HC-CAU. Each cluster represents altered chemical class of metabolites (*p* <0.05). Cluster sizes represent the total number of metabolites. Cluster color represents the directionality of metabolite differences: red – higher in HC-HIS; blue – lower in HC-HIS. Colors in between refer to mixed population of metabolites manifesting both higher and lower levels in HC-HIS when compared to the HC-CAU. The *x*-axis represents the cluster order on the chemical similarity tree. The plot *y*-axis shows chemical enrichment *p*-values calculated using Kolmogorov–Smirnov test. Only clusters with *p* <0.05 are shown. FDR-adjustment *q* =0.2, and clusters with FDR-adjusted *p* ≥0.2 are shown in gray. (*n*, HC-HIS =14, HC-CAU =8) The detailed ChemRICH results are shown in (Table S4). AC, Acylcarnitines; CAU, White Caucasian; DG, Diglycerides; Ether-PC, Ether-linked phospholipids; HIS, Hispanic; MUFA, Monounsaturated fatty acid; PUFA, Polyunsaturated fatty acids; TG, Triglycerides.

### 4. Ethnicity-specific alteration in hepatic FFAs profile characterizes NASH

To examine differences in hepatic metabolomic profile associated with the progression to NASH, we stratified NAFL group into 0-NASH and NASH. Due to the limited sample size and breadth of current analysis, we first examined NASH-dependent changes in each ethnicity by chemical classes. Next, to examine ethnicity-specific metabolomic alterations in NASH, ANCOVA was performed with interaction (ethnicity x NASH) followed by variable clustering for data reduction, given the strong class-specific changes observed, the small sample size and the exploratory nature of this pilot.

Upon examining liver metabolomic profile, ChemRich analysis (Fig. 4) showed that NASH was associated with higher unsaturated FFAs in HIS, driven by heptadecenoic acid (17:1n7); lower unsaturated PCs concurrent with higher levels in unsaturated LPCs; lower levels of unsaturated PEs with an elevation in many ether-linked PLs; lower ACs with higher TGs and DGs. To a lesser impact, HIS-NASH showed alteration in organic compounds, including lower level of several amino acids and related derivatives and in purine nucleosides and adenosine (Table S6). NASH in CAU showed higher SFAs, mainly capric acid (10:0) and lauric acid (12:0); lower levels of many unsaturated FFAs, PE, phosphatidylserines and in organic compounds including trimethyl ammonium compounds and dipeptides (Table S7).

**Fig 4.**
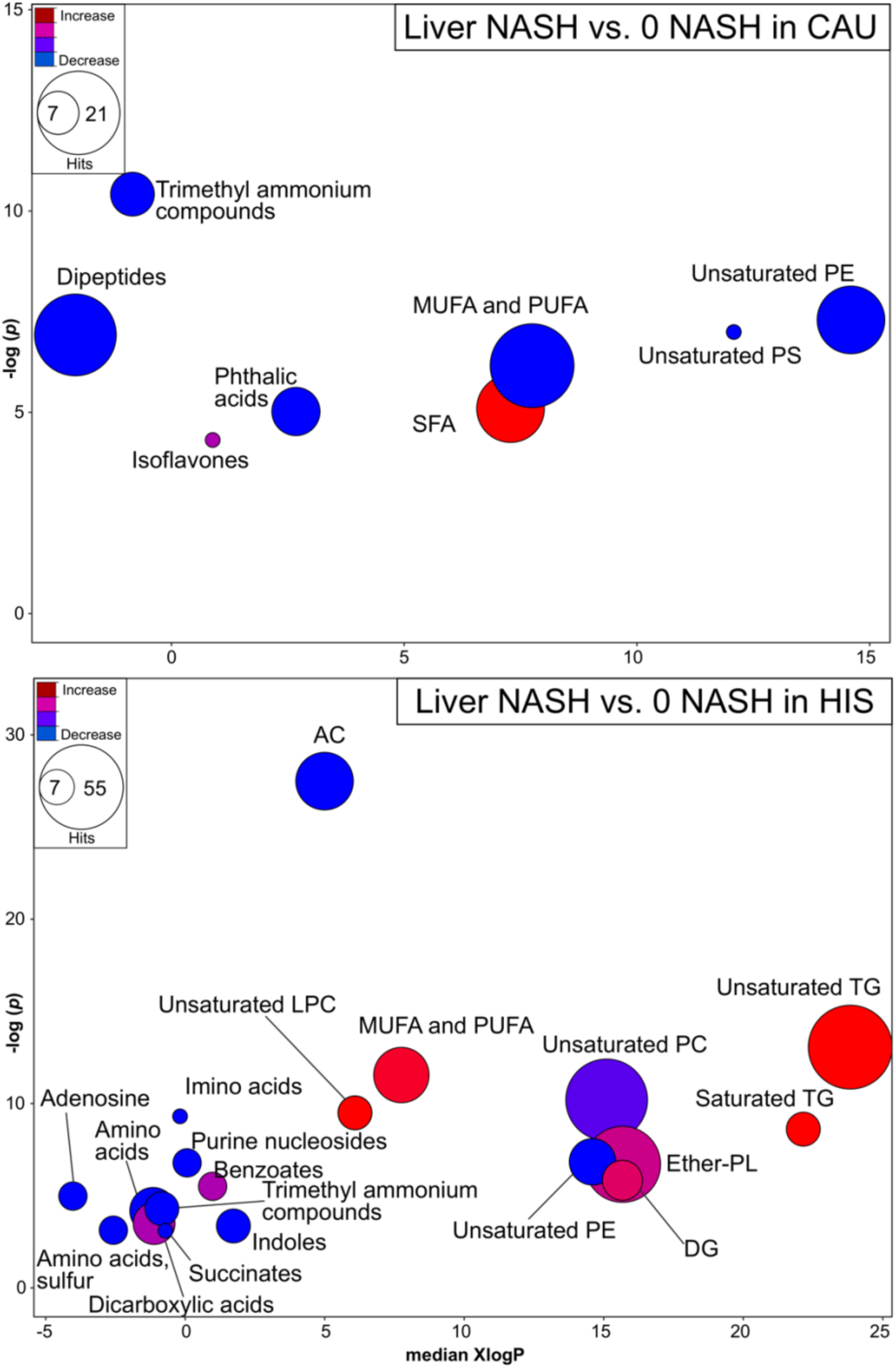
Liver metabolites altered by chemical class in NASH, compared to NASH -free subjects, in both ethnicities. Chemical similarity enrichment analysis (ChemRICH) illustrating hepatic metabolite class altered in NASH compared to 0-NASH in both ethnicities. ChemRICH enrichment statistics plot, illustrating liver alterations in NASH vs. 0-NASH in CAU (top panel) and HIS (bottom panel). Each cluster represents altered chemical class of metabolites (*p* <0.05). Cluster sizes represent the total number of metabolites. Cluster’s color represents the directionality of metabolite differences: red – higher in NASH; blue – lower in NASH. Colors in between refer to mixed population of metabolites manifesting both higher and lower levels in NASH when compared to the 0-NASH. The *x*-axis represents the cluster order on the chemical similarity tree. The plot *y*-axis shows chemical enrichment *p*-values calculated using Kolmogorov–Smirnov test. Only clusters with *p* <0.05 are shown. FDR-adjustment *q* =0.2, and clusters with FDR-adjusted *p* ≥0.2 are shown in gray. (*n*, 0 NASH-HIS =4, NASH-HIS =3; 0 NASH-CAU =5, NASH-CAU =5). The detailed ChemRICH results are shown in (Table S6 and S7). AC, Acylcarnitines; CAU, White Caucasian; Cer, Ceramides; CE, Cholesteryl ester; DG, Diglycerides; Ether-PL, Ether-linked phospholipids; HIS, Hispanic; LPC, Lysophosphatidylethanolamines; MUFA, Monounsaturated fatty acid; NASH, Nonalcoholic steatohepatitis; 0-NASH, Nonalcoholic steatohepatitis-free; PC, Phosphatidylecholine; PE, Phosphatidylethanolamine; PUFA, Polyunsaturated fatty acids; SFA, Saturated fatty acids; TG, Triglycerides.

ANCOVA showed 26 liver metabolites, (2.8%) of detected metabolites being altered at a *p* <0.05 between ethnicities in case of NASH with an additional 33 metabolites showing a *p*-value between 0.05 and <0.1. However, these did not survive FDR-correction. Regardless, we proceeded with the characterization of the composition of these metabolites and their distribution between the groups.

Variable clustering performed on metabolites with *p* <0.05 for the NASH effect within ethnicity groups and with *p* <0.05 for the ethnicity x NASH interaction collapsed the data into 32 cluster components. Comparison of cluster components between ethnicities (Fig. 5) suggested that with NASH, clusters 3, 4, 6, 7, 8, 16, 18, 22, 23, 24 were differentially altered with tendencies shown for cluster 2, 9, 13, 14, 26. Cluster descriptions, correlations, means and *p*-values for cluster components and individual metabolites are detailed in Table S8.

**Fig 5.**
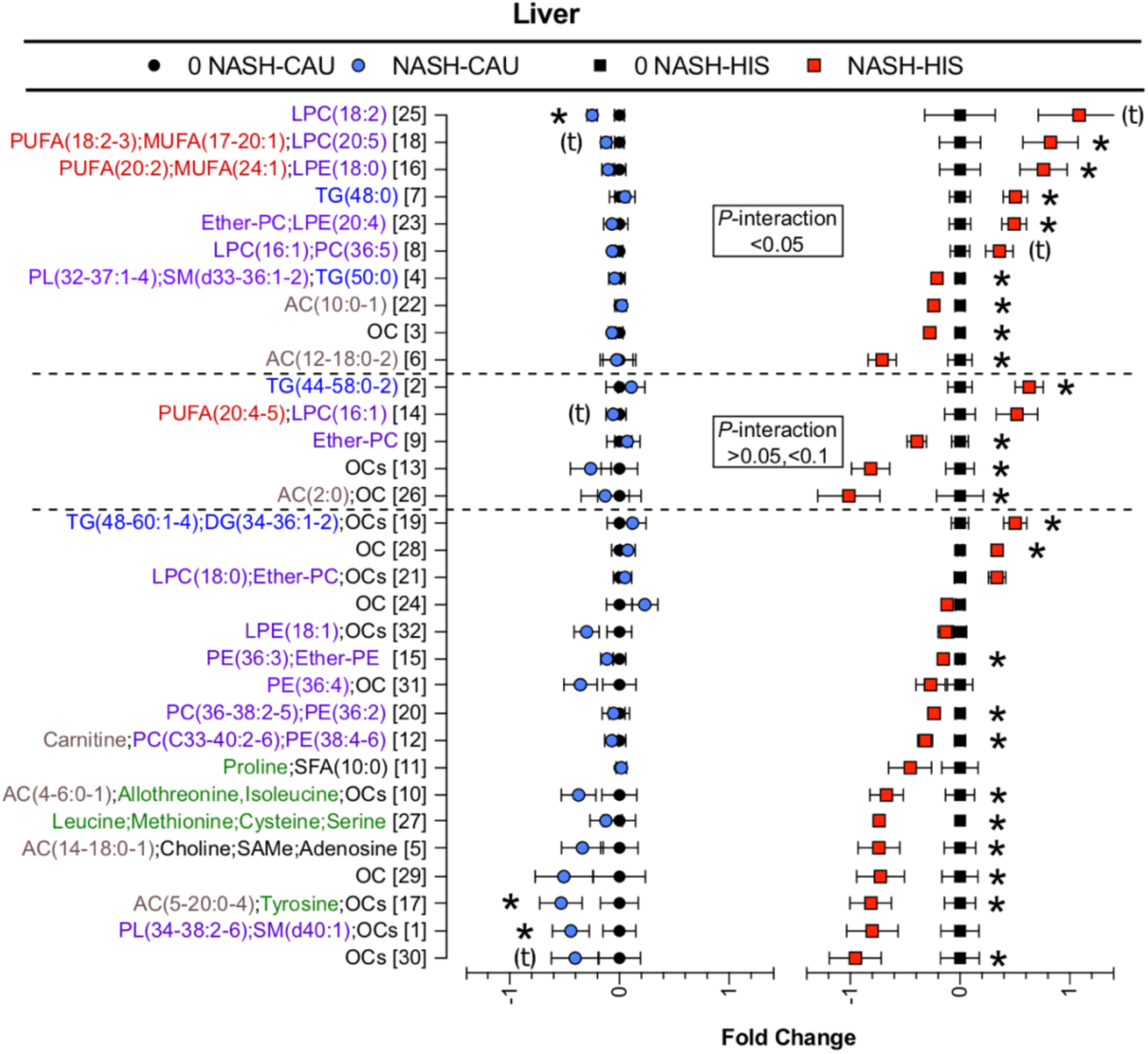
Ethnicity-related variations in liver metabolomic profile in NASH vs. 0-NASH. Results of *t*-test and ANCOVA (ethnicity x NASH) interaction comparison performed on plasma variable cluster components between NASH and 0-NASH in both ethnicities. Clustering was performed on metabolites affected by NASH within and between ethnicity groups ethnicity (*p* <0.05). Data are presented as the fold change from 0-NASH; error bars represent standard error. Metabolite clusters are labeled by a number and a description of representative metabolite. For complex lipids, lipid class is followed by number of (carbons, double bounds) of the fatty acyl moiety. Clusters with interaction (*p* <0.05) or tendency for interaction (*p* =0.05 to < 0.1) are marked with the dashed line. Clusters affected by NASH within ethnicity (*p* <0.05) are denoted with (*); clusters with tendency (*p* =0.05 to < 0.1) are denoted with (t). (*n*, 0 NASH-HIS =4, NASH-HIS =3; 0 NASH-CAU =5, NASH-CAU =5). The details on metabolites and clusters components are shown in (Table S8). AC, Acylcarnitines; CAU, White Caucasian; DG, diglycerides; Ether-PC, Ether-linked phosphatidylcholines; Ether-PE, Ether-linked phosphatidylethanolamines; HIS, Hispanic; MUFA, Monounsaturated fatty acid; NASH, Nonalcoholic steatohepatitis; 0-NASH, Nonalcoholic steatohepatitis-free; OC, Organic compounds; PC, Phosphatidylcholines; PE, Phosphatidylethanolamine; PL, Phospholipids; PUFA, Polyunsaturated fatty acids; NAFL, Steatosis; SAMe, S-Adenosyl-L-methionine; SFA, Saturated fatty acids; SM, sphingomyelins; TG, triglycerides.

NASH appeared associated with differential changes in the hepatic FFA profiles between ethnicities. Among the altered clusters was cluster 18 (*p* <0.05 for interaction of ethnicity x NASH). This cluster is composed of the MUFAs, heptadecenoic acid (17:1n7), oleic acid (18:1n9) and eicosenoic acid (20:1n9); and the PUFAs, linoleic acid (18:2n6), α-linolenic acid (18:3n3). These FFAs were higher with NASH in HIS with an opposite trend seen in CAU. Similar trends were observed for arachidonic acid (20:4n6) and eicosapentanoic acid (20:5n3) (cluster 14), however, this cluster showed tendency for interaction (ethnicity x NASH). The MUFAs and PUFAs in cluster 14 and 18 correlated with LPC(16:1) and LPC(20:5).

Hepatic LPC and LPE profiles appeared to be differentially altered between ethnicities in NASH with higher levels seen in HIS and lower levels in CAU and interaction shown for LPC(16-20:1-5) (clusters 18, 8, 25) and LPE(18-20:0-4) (cluster 16 and 23), and with tendency shown for LPC(16-22:1-6) (cluster 2, 14). For both ethnicities, NASH had lower levels of almost all hepatic PCs and PEs. This trend differed between ethnicities in HIS for PC(32-37:1-3) and PE(34-36:2-4) (cluster 4), with apparent alterations in some ether-linked PLs (cluster 9 and 23) and SM(d33-36:1-2) (cluster 4).

There also appeared to be a lower level of hepatic ACs in NASH cases. However, a differentially lower ACs with interaction (ethnicity x NASH) was shown in HIS for AC(10-18:0-2) (cluster 22, 6) with tendency for AC(2:0) (cluster 26). In both ethnicities, NASH showed higher hepatic TGs, with differentially higher (*p* <0.05 in NASH between ethnicities) seen for TG(44-58:0-2) (cluster 2) and tendency shown for TG(48-50:0) (cluster 7, 4) observed in HIS. Some organic compounds of unspecific classification (cluster 3, 13, 14, 8) were also differentially altered in NASH between ethnicities.

Collectively, these findings suggest ethnicity-related differences in hepatic metabolomic profile with the progression to NASH, with markedly increased hepatic FFAs, TGs; lower levels of ACs and changes in several PLs seen in NASH-HIS. A heatmap illustrating variations in plasma and liver FFAs profile with NASH is shown in Fig. 6. Additionally, we performed pathway analysis using hepatic metabolites seen to be different between ethnicity group in NASH (raw *p* <0.05). These results support our cluster analysis with pathways related to unsaturated fatty acid metabolism found altered (FDR-adjusted (*p* <0.2), reflecting alteration in linoleic acid (18:2n6), eicosapentaenoic acid (20:5), oleic acid (18:1n9) and several PLs (Fig 7 and Table S9).

**Fig 6.**
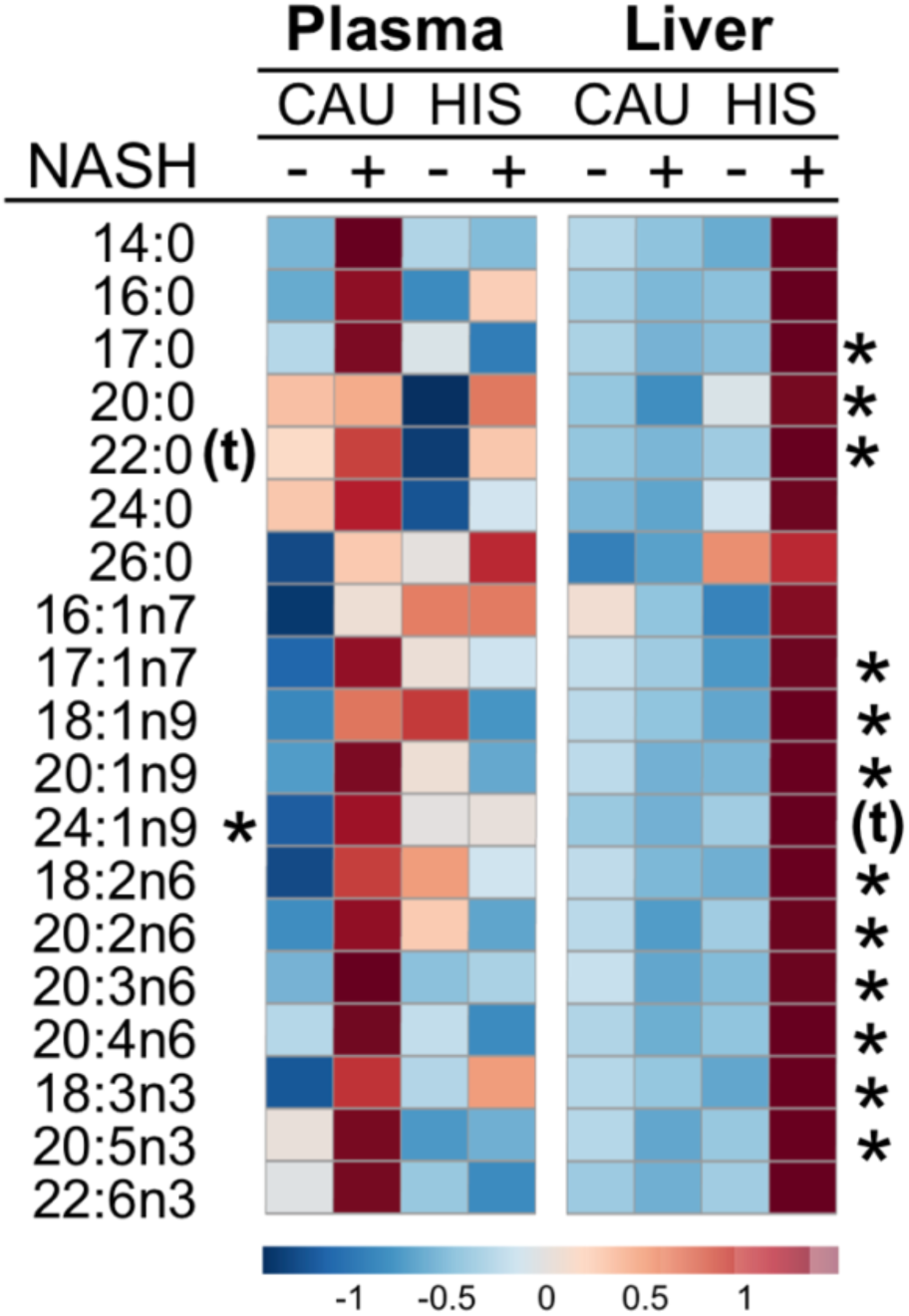
Heat-map for plasma and liver free fatty acids with fold change and *p*-values in NASH vs. 0 NASH in both ethnicities. Group means are shown with normalized data converted to z-scores. Fatty acids with interaction (*p* <0.05) by ethnicity in NASH are noted by (*), and with tendency for interaction (*p* =0.05 to <0.1) are denoted with denoted with (*t*). Mean direction of change is indicated by color and intensity, with red representing increased values, and blue representing decreased values. CAU, White Caucasian; HIS, Hispanics; NASH, Nonalcoholic steatohepatitis.

**Fig 7.**
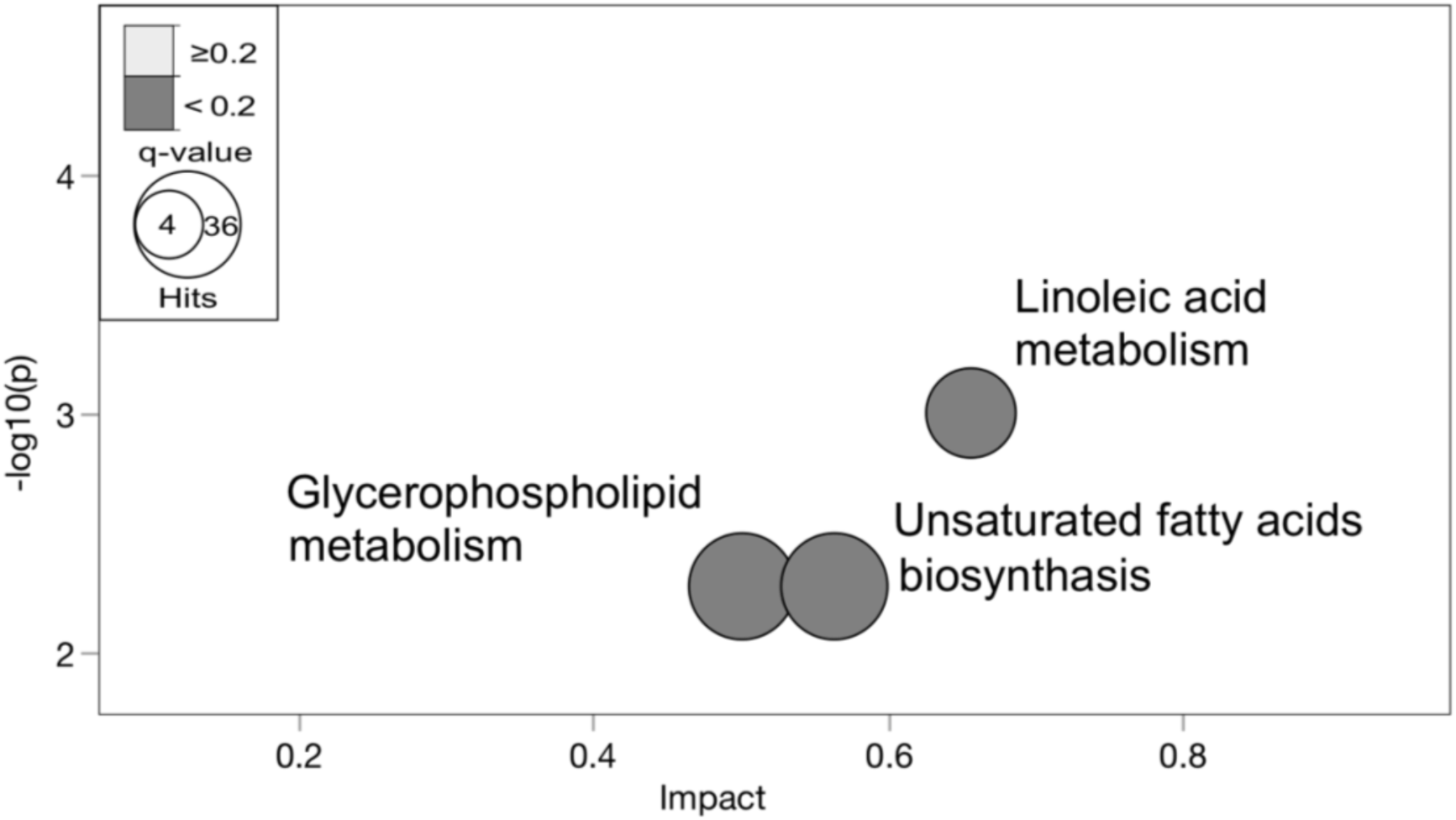
Hepatic metabolic pathways differentially altered with NAHS between ethnicities. Metabolites with differential alterations between ethnicity in NASH (*p* <0.05) were compared against pathway-associated metabolites sets from Kyoto Encyclopedia of Genes and Genomes (KEGG) [46]. Metabolic pathways significantly altered are shown as nodes. The (*y*-axis) represents the *p*-values as determined by Fisher’s Exact test. The (*x-*axis) represents the impact of pathways as determined by the relative betweenness centrality-topology analysis. The size of the node represents the total hit number of hits. (*n*, 0 NASH-HIS=4, NASH-HIS=3; 0 NASH-CAU=5, NASH-CAU=5). Detailed pathway analysis statistics are shown in (Table S9).

### 5. Ethnicity-specific alteration in plasma metabolites characterizing NASH

When examining NASH-dependent changes by chemical classes in plasma and in comparison to 0-NASH groups, ChemRich analysis (Fig. 8) showed that NASH-HIS had higher unsaturated TGs and SMs (Table S10) and NASH-CAU had higher Cers and SMs (Table S11).

**Fig 8.**
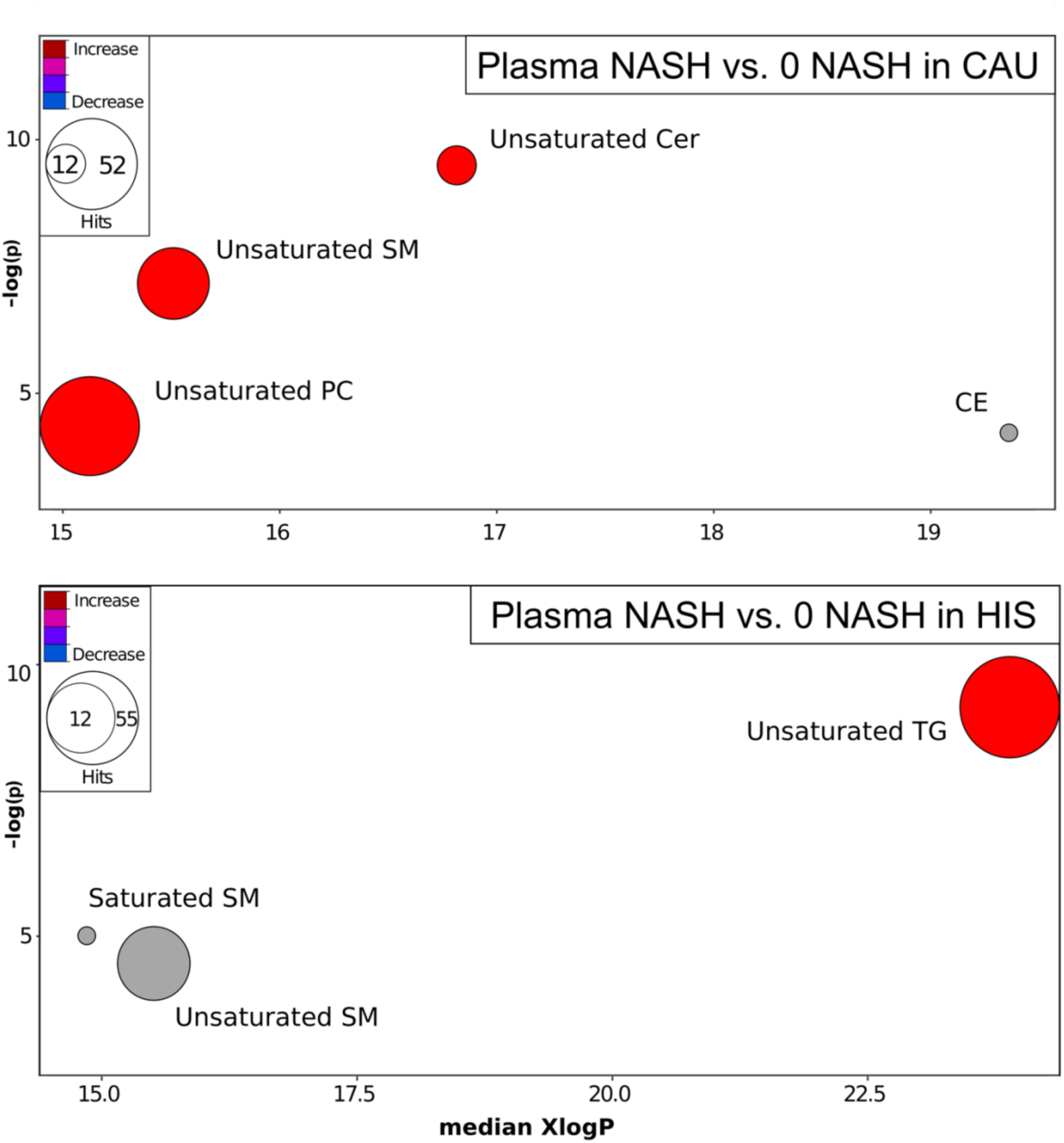
Plasma metabolites altered by chemical class in NASH, compared to NASH -free subjects, in both ethnicities. Chemical Similarity Enrichment Analysis (ChemRICH) of illustrating plasma metabolite class altered in NASH compared to 0-NASH in both ethnicities. ChemRICH enrichment statistics plot, illustrating plasma alterations in NASH vs. 0-NASH in CAU (top panel) and HIS (bottom panel). Each cluster represents altered chemical class of metabolites (*p* <0.05). Cluster sizes represent the total number of metabolites. Cluster’s color represents the directionality of metabolite differences: red – higher in NASH; blue – lower in NASH. Colors in between refer to mixed population of metabolites manifesting both higher and lower levels in NASH when compared to the 0-NASH. The *x*-axis represents the cluster order on the chemical similarity tree. The plot *y*-axis shows chemical enrichment *p*-values calculated using Kolmogorov–Smirnov test. Only clusters with *p* <0.05 are shown. FDR-adjustment *q*=0.2, and clusters with FDR-adjusted *p* ≥0.2 are shown in gray. (*n*, 0 NASH-HIS=4, NASH-HIS=3; 0 NASH-CAU=5, NASH-CAU=3). The detailed ChemRICH results are shown in (Table S10 and S11). CAU, White Caucasian; Cer, Ceramides; CE, Cholesteryl ester; HIS, Hispanic; SM, Sphingomyelins; NASH, Nonalcoholic steatohepatitis; PC, Phosphatidylcholine; TG, Triglycerides.

To examine ethnicity-specific metabolomic alterations in NASH, we performed ANCOVA with interaction (ethnicity x NASH) followed by variable clustering for data reduction. Results showed 18 (1.9%) of detected plasma metabolites appeared different at a *p* <0.05 in NASH between ethnicities, with 13 additional metabolites showing a *p*-value between 0.05 and <0.1. These differences did not survive FDR-correction. Cluster analysis reduced data dimensionality into 25 clusters. Comparison of cluster components (Fig. 9) showed that clusters 4, 5, 17, 20, 25 were differentially altered with NASH between ethnicities with tendencies shown for cluster 7, 11, 12, 13, 16. Cluster descriptions, correlations, means and *p*-values for cluster components and individual metabolites are detailed in Table S12.

**Fig 9.**
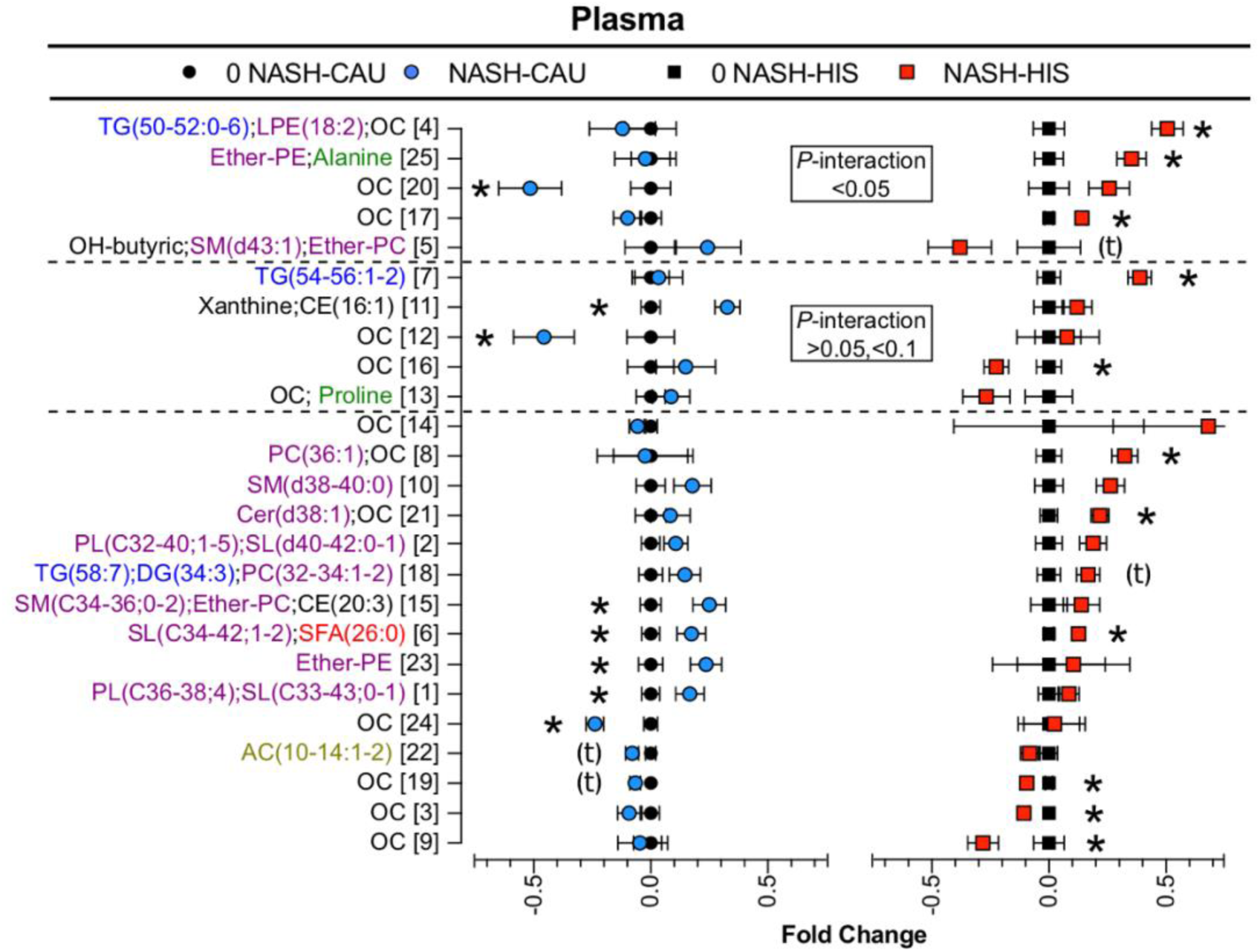
Ethnicity-related differences in plasma metabolomic profile in NASH vs. 0-NASH. Results of *t*-test and ANCOVA (ethnicity x NASH) interaction comparison performed on plasma variable cluster components between NASH and 0-NASH in both ethnicities. Clustering was performed on metabolites affected by NASH within and between ethnicity groups ethnicity (*p* <0.05). Data are presented as the fold change from 0-NASH; error bars represent standard error. Metabolite clusters are labeled by a number and a description of representative metabolite. For complex lipids, lipid class is followed by number of (carbons, double bounds) of the fatty acyl moiety. Clusters with interaction (*p* <0.05) or tendency for interaction (*p*=0.05 to <0.1) are marked with the dashed line. Clusters affected by NASH within ethnicity (*p* <0.05) are denoted with (*); clusters with tendency (*p*=0.05 to <0.1) are denoted with (*t*). (*n*, 0 NASH-HIS=4, NASH-HIS=3; 0 NASH-CAU=5, NASH-CAU=3). The details on metabolites and clusters components are shown in (Table S12). AC, Acylcarnitines; CAU, White Caucasian; CE, cholesteryl ester; DG, diglycerides; Ether-PC, Ether-linked phosphatidylcholines; Ether-PE, Ether-linked phosphatidylethanolamines; HIS, Hispanic; MUFA, Monounsaturated fatty acid; NASH, Nonalcoholic steatohepatitis; 0-NASH, Nonalcoholic steatohepatitis-free; OC, Organic compounds; PC, Phosphatidylcholines; PL, Phospholipids; PUFA, Polyunsaturated fatty acids; NAFL, Steatosis; SFA, Saturated fatty acids; SM, sphingomyelins; TG, triglycerides.

With NASH in HIS, the plasma TGs profile was differentially altered (*p* <0.05 for ethnicity x NASH interaction) with higher levels of TG(50-52:0-6) (cluster 4) and tendency for TG(54-56:1-2) (cluster 7). Compared to respective 0-NASH, plasma levels of CEs, PCs, and PEs were higher in NASH. Ethnicity-specific differences included CE(16:1) (cluster 11), which was higher in CAU; LPE(18:2) (cluster 4) and ether-linked PLs (cluster 5 and 25), which were higher in HIS.

Differences in FFA profiles were not observed in plasma between NASH and 0-NASH. Both ethnicities showed lower plasma AC(10-14:1-2) (cluster 22) with NASH, however, HIS had a differentially lower (*p* <0.05 for ethnicity x NASH interaction) 3-hydroxybutyric acid (cluster 5). NASH was also associated with specific plasma amino acid profile with interaction (ethnicity x NASH) seen in HIS with higher alanine (cluster 25). Also, differential or with tendency for interaction are alterations in some organic compounds (cluster 4, 7, 12, 13, 15, 16, 17, 20 and 25).

Together, these findings reveal that ethnicity-related differences associated with NASH in plasma mainly affected TGs and some PLs in HIS, with altered organic compounds seen in both ethnicities.

### 6. Correlations of metabolomic profiles with liver histology

Correlation analyses were performed on hepatic metabolites with steatosis and lobular inflammation, two histological features relevant to NASH diagnosis. In NASH-CAU, lobular inflammation correlated positively with metabolites including the eicosanoid, 20-hydroxyarachidonic acid; and negatively with MUFA(16-18:1); TG(50:0) (*r* =-0.6, raw *p* <0.05). Hepatic steatosis correlated negatively with metabolites including MUFA(20-24:1); arachidonic acid (20:4n6); choline and S-Adenosyl-L-homocysteine. It correlated positively some SFA(9-12:0), TG(49:0) (*r* >-0.6, raw *p* <0.05).

In NASH-HIS, hepatic inflammation correlated negatively with metabolites including AC(5-20:0-4); purine related metabolites, adenosine, inosine, hypoxanthine; SAMe; Cer(d40:1-2), and PC(33-40:1-7). It correlated positively with DG(34-36:1-3); TG(44-60:0-4); the MUFA(17-24:1); PUFA, α-linolenic acid (18:3n3), linoleic acid (18:2n6), eicosadienoic acid (20:2n6); and LPC(18-20:0-5) (*r* >0.7, raw *p* <0.05). These metabolites retained significance after FDR-adjustments. Hepatic steatosis correlated positively with TG(48-58:2-8) and negatively with some organic compounds (*r* >0.7, raw *p* <0.05) (Table S13). Conversely, hepatic steatosis and lobular inflammation scores correlated with many plasma metabolites, but these did not pass FDR-adjustment (Table S14).

## Discussion

Hispanics are impacted with higher prevalence and progression rate of NAFLD [6–11]. However, the biological background for this disparity is not clear. To investigate metabolic variations associated with NAFL in these groups, we examined plasma and liver metabolomic profiles with respect to ethnicity in a group of HIS and CAU subjects with obesity and liver biopsy-characterized NAFL. Results revealed several ethnicity-related metabolomic distinctions and the major findings are: 1) In NAFL, as compared to HC, the plasma FFAs and ACs profile are differentially altered between ethnicities; 2) HIS present with signs of metabolic perturbation independent of obesity; 3) In NASH, comparing to BMI and histology-matched NASH-free subjects, the hepatic profile was differentially altered between ethnicities with HIS showing higher abundance FFAs and lipotoxic lipids.

In NAFLD, peripheral insulin resistance contributes to elevated plasma FFAs as the insulin-mediated suppression of adipose tissue lipolysis is impaired [47, 48]. In the current study, NAFL groups were obese with features of metabolic syndrome. However, our results show discrepancies in plasma profiles between ethnicities. In NAFL-CAU, there were higher plasma FFAs, as compared to HC. This is consistent with insulin resistance, increased adipose tissue efflux and is in line with previous lipidomic analysis in NAFLD subject [24, 29, 49]. NAFLD is also characterized by impaired metabolic flexibility and plasma ACs are reported to be altered [25, 50]. In NAFL-CAU, we confirmed an elevation in many ACs species, including short chain AC(2-5:0) and long chain AC(10-14:0-2). AC(3-5) are derived from amino acids, even-chain AC(4-20) reflect incomplete fatty acid β-oxidation and AC(2:0) is the product of complete β-oxidation and may originate from glucose metabolism [51]. Altered plasma ACs profile indicates dysregulations in substrate oxidation across all tissues.

Unexpectedly, elevations in plasma FFAs and in ACs profiles were not marked in NAFL-HIS, which can be explained by the higher ACs and unsaturated fatty acids seen in lean and healthy HIS, as compared to CAU counterparts. We also observed higher plasma TGs in HC-HIS, a feature often associated with insulin resistance and obesity [52]. Hypertriglyceridemia is reported to be more frequent in overweight and obese HIS, compared to other ethnicities [53–56]. However, our findings indicate that this derangement along with a higher FFAs, ACs, altered PLs and amino acids profiles are independent of obesity and suggest early signs of metabolic perturbation in HC-HIS. Given that NAFLD results from a complex interplay between environmental factors, host genetics, intestinal dysbiosis and comorbidities [13], it is possible that such metabolic derangements increase the predisposition of HIS to NAFLD and/or its associated risk factors and are worth further investigation.

Our findings also show lower ether-linked PLs in NAFL-CAU. Although these differences were not evident in NAFL-HIS, a lower abundance of many ether-linked PLs were observed in HC-HIS, compared to HC-CAU. In NAFLD, a reduction in ether-linked PLs correlated with disease severity [29]. Beside a role in maintaining bio membrane fluidity and integrity, ether-linked PLs are thought to ameliorate NAFLD by modulating fatty acid oxidation and an antioxidant capacity [57, 58].

With the progression from NAFL to NASH, our findings show differences in hepatic lipidomic profile between ethnicities, despite comparable NAS scores. Remarkably, NASH-HIS subjects had higher levels of hepatic FFAs with ethnicity interaction (*p* <0.05) shown for the MUFA heptadecenoic acid (17:1n), oleic acid (18:1n9), eicosenoic acid (20:1n9); and in the PUFA, α-linolenic acid (18:3n3), linoleic acid (18:2n6). Results from hepatic pathway analysis corroborate our metabolomic findings and show unsaturated fatty acid metabolism pathways differentially altered with NASH between ethnicities. These ethnicity-related alterations in hepatic FFAs suggest variations in processes including hepatic uptake of FFAs, DNL, PLs turnover and hepatic unsaturated fatty acid metabolism, which are worth further examination.

Upregulated hepatic FFA clearance may explain the observed elevation of hepatic FFAs in NASH-HIS. In fact, tracer studies in NAFLD show that 60% of hepatic lipid content originates from adipose tissue lipolysis, indicating a major contribution of the liver’s uptake to hepatic FFAs profile [59]. In support, in both animals and humans, the expression and membrane localization for fatty acid translocase CD36 are increased in NASH [60].

Hepatic TGs accumulation is thought to be a protective adaptive mechanism from the lipotoxic effect of FFAs [61–63]. However, this notion is questioned as in subjects with early NAFL and no histological evidence of inflammation, the expression of several hepatic inflammatory genes were upregulated [64]. In NASH, DNL was reported to be increased in comparison to NAFL [65]. This process generates SFA(14-16:0), which are converted to MUFA via stearoyl-CoA desaturase (SCD1, or Δ-9 desaturase) and further esterified to form DGs and TGs [66]. In our study, NASH in both ethnicities showed no differences in the estimated hepatic Δ-9 desaturase index (results not shown). NASH also had higher level of TGs, but more profound in HIS with a trend of higher DGs. The higher levels of TGs, DGs and MUFA observed in NAHS-HIS suggest increased DNL. In NASH-HIS, lobular inflammation correlated positively with TG(44-60:0-4) (*r* >0.7, *p* <0.05), DG(34-36:1-3) (*r* >0.7, *p* <0.000) and the MUFAs, heptadecenoic acid (17:1n7), oleic acid (18:1n9), eicosenoic acid (20:1n9), and nervonic acid (24:1n9) (*r* >0.8, *p* <0.00) suggesting an involvement of TGs, DGs, MUFAs with NASH in HIS. Interestingly, in NASH-CAU a negative correlation (*r* >-0.6, *p* <0.05) was observed between hepatocellular inflammation and heptadecenoic acid (17:1n7), oleic acid (18:1n9), with tendency shown for palmitoleic acid (16:1n7). While, when tested individually, MUFAs are shown to be less lipotoxic compared than SFAs, it has been shown that treatment of HepG2 cells and human primary hepatocytes with a mixture of five SFAs and MUFAs as observed in NASH, exhibited higher toxicity compared to the mixture reported in normal liver and NAFL [21, 24]. Therefore, the effect of MUFAs may be dependent factors including proportion and composition. Also in our samples, plasma TGs were differentially higher in NASH-HIS, which suggests upregulated hepatic export or impaired peripheral clearance.

An upregulated PL turnover can contribute to elevated unsaturated fatty acids. In our results, the strong correlation found between the altered MUFAs, PUFAs and many lysophospholipid species (Table S8) suggests a shared biochemical pathway. As part of hepatic lipid remodeling, phospholipase A2 catalyzes the breakdown of membrane PL to form lysophospholipids and release unsaturated fatty acids. In NASH, the transcript level of cytosolic phospholipases A2 (*cPLA2*) is upregulated [67]. Also, NASH is markedly lower in hepatic PCs content and higher in LPCs [24, 28, 68]. Our results are in line with the decreased hepatic PCs and PEs profile seen in NASH in both ethnicities, with more marked reductions seen in NAHS-HIS. However, the hepatic LPC and LPE profile showed differential alterations by ethnicity with higher levels observed in HIS and lower levels in CAU. Together, the higher abundance of hepatic PUFA, lower level of PC and higher LPC seen in NASH-HIS support an upregulated hepatic PL turnover. The depletion in PC or alteration in PC to PE ratio disrupts the function and integrity of cellular, mitochondrial and lipid droplets bio-membranes and results in cellular stress, apoptosis and inflammation [69]. *In vitro* evidence shows a lipotoxic effect of LPC by altering mitochondrial function and activating apoptosis [23, 70, 71]. In HIS-NASH, lobular inflammation correlated positively with LPC(16-20:0-5) and the PUFA, linoleic acid (18:2n6), α-linolenic acid (18:3n3) and eicosadienoic acid (20:2n6); and negatively with PC(32-40:1-7) and PE(34-38:2-6) (*r* >0.8, *p* <0.006), which are also in support of a role of upregulated PLs turnover in NASH pathogenesis.

A decrease in hepatic fatty acid desaturase-1(FADS1, Δ-5 desaturase) is reported with NASH [24]. This is thought to induce preferential increase in n6 PUFAs flux at the expense of n3 PUFAs. Accordingly, NAFLD progression is characterized by de-enrichment of long-chain PUFAs with a shift towards a higher n6 to n3 PUFAs ratio seen across multiple lipid classes [27, 28]. In the current study, we estimated Δ-5 desaturase and the relative ratio of n6 to n3 PUFAs from hepatic FFAs and results show no differences with NASH for both ethnicities (not shown).

Also, the progression to NASH was associated with reduction in most hepatic ACs in both ethnicities with differentially lower AC(12-18: 0-2) seen in HIS. We also found the hepatic AC(5-18:0-4) correlated negatively with lobular inflammation in NASH-HIS. In plasma, 3-hydroxybutyric acid was found differentially reduced in NASH-HIS. 3-hydroxybutyric (or β-hydroxybutyrate) is a liver-produced ketone body from acetyl-CoA derived from mitochondrial β-oxidation of fatty acid [72]. Together, these findings suggest a role of mitochondrial β-oxidation dysfunction in both ethnicities with NASH.

Alterations in hepatic amino acid, choline, methionine and SAMe profiles were associated with NASH but not different between ethnicities. The dysregulations in hepatic unsaturated fatty acid metabolism associated with NASH progression were not reflected in plasma. This, along with the limited correlations of plasma metabolomic profile with liver histological scores, leads us to conclude plasma profile is not an optimal predictor of hepatic histological differences relevant to NASH.

Our data may be used for hypotheses generation. We did not account for diet and treatment with lipid-lowering drugs, which are commonly prescribed in individuals with medically complicated obesity. Since performing a liver biopsy without clinical indication is ethically questionable, we could not confirm that our HC subjects are free from liver pathology. It might have been interesting to genotype our subjects for *PNPLA3* as the G allele is robustly associated with NAFLD and the homozygous rs738409 [G] variant presents with high frequency in HIS [73–75]. Due to sample size, cross-sectional and proof-of-concept nature, our findings need to be confirmed in larger and mechanistic settings. In particular, based on selected metabolites with ethnicity interaction (*p* <0.05) (Table S15), the minimum sample size required to verify plasma findings in lean healthy subjects is 45 subjects per group. To verify alterations in hepatic lipidomic profile, a minimum of 24 and 12 subjects would be required for each arm in CAU and HIS, respectively.

To our knowledge, this is the first metabolomic profiling addressing ethnicity in NAFLD. Our NAFL patients are biopsy-characterized with comparable BMI, clinical and histological presentations; with almost all plasma and liver samples obtained from the same subjects. Our findings provide preliminary evidence supporting ethnicity-related variations in NAFLD pathogenesis and highlight signs of metabolic perturbations in HIS independent of obesity and other components of metabolic syndrome. With the progression to NASH, our data suggest ethnicity-related alterations in hepatic unsaturated fatty acids metabolism and points toward potential involvement of lipotoxicity in its mechanisms. We postulate that such alterations predispose HIS to higher rate of NAFLD and/or add a “hit” component when coexisting with metabolic syndrome to act as a driver for the advanced NASH presentation seen in HIS.

## Data Availability

The study details are available on The Metabolomics Workbench (http://www.metabolomicsworkbench.org), ID number (ST000977).

## Abbreviations

AC: Acylcarnitine
CAU: White Caucasians
CE: Cholesteryl ester
Cer: Ceramide
cPLA2: Cytosolic phospholipases A2
CSH-QTOF MS: Charged surface hybrid column/quadrupole time of flight mass spectrometer
DG: Diglyceride
DNL: *De novo* lipogenesis
FDR: False discovery rate
FFA: Free fatty acid
GC-TOF MS: Gas chromatography/time-of-flight mass spectrometry
HC: Healthy control
HILIC-QTOF MS/MS: Hydrophilic interaction liquid chromatography/quadrupole time of flight mass spectrometer
HIS: Hispanics
KEGG: Kyoto Encyclopedia of Genes and Genomes
LPC: Lysophosphatidylcholine
LPE: Lysophosphatidylethanolamine
MUFA: Monounsaturated fatty acid
NAFL: Steatosis
NAFLD: Nonalcoholic fatty liver disease
NAS: The NAFLD Activity Score
NASH: Nonalcoholic steatohepatitis
NASH-CRN: The NASH Clinical Research Network
PC: Phosphatidylcholine
PCA: Principal component analysis
PE: Phosphatidylethanolamine
PL: Phospholipid
PLS-DA: Partial least square-discriminant analysis
PUFA: Polyunsaturated fatty acid
SAMe: S-Adenosyl-L-methionine
SFA: Saturated fatty acid
SM: Sphingomyelin
TG: Triglyceride
TMAO: Trimethylamine N-oxide
VIP: Variable importance in projection
0-NASH: Nonalcoholic steatohepatitis-free
Δ-5 desaturase: Fatty acid desaturase-1
Δ-9 desaturase: Stearoyl-CoA desaturase.

## Author Contributions

Original draft writing-T.A.M.; Review & Editing-T.A.M., V.M., K.B., J.W.N., K.L.S, P.J.H., M.A., C.L.B, S.S., K.M., Y.Y.W.; Formal Analysis-K.B and T.A.M.; Visualization-T.A.M. and K.B ; Methodology-V.M., O.F., D.K, T.A.M, K.M, Y.Y.W.; Investigation-T.A.M, K.M, Y.Y.W.; Resources-V.M., O.F., C.L.B, S.S., K.L.S., P.J.H., M.A.; Funding Acquisition-V.M., P.J.H.

## Funding

This work was supported by The National Institutes of Health (NIH)-West Coast Metabolomic Center (WCMC) grant number DK104770 (to V.M.), NIH-National Heart, Lung and Blood Institute 1R01 HL09133 (to P.H.) and 1R01 HL107256 (to P.H.). The content is solely the responsibility of the authors and does not necessarily represent the official views of the NIH and WCMC.

## Conflict of interest

The authors declare no conflict of interest and nothing to disclose.

## Supplemental figures and tables legend

**Fig S1. Natural clustering of study groups illustrating variations in plasma metabolomic profile.** Cluster components of metabolites with potential ethnicity interaction (*p* <0.1) were projected onto principal component analysis (PCA). The PCA clustering is presented as **a)** loading plot showing all subjects in NAFL and HC groups; **b)** score plot showing cluster components (numbered 1 to 20). The PCA component number is shown with correspondent explained variance in brackets. CAU, White Caucasian; HC, Healthy control; HIS, Hispanic.

**Table S1.** ChemRich enrichment statistics for plasma comparisons of obese with NAFL vs. lean healthy control in White Caucasian. Details are shown for clusters with *p* <0.05. Clusters *p*-values was calculated using Kolmogorov–Smirnov test. FDR-adjusted *q* =0.2, with (1) FDR-adjusted *p* <0.2; (0) FDR-adjusted *p* ≥0.2.

**Table S2.** ChemRich enrichment statistics for plasma comparisons of obese with NAFL vs. lean healthy control in Hispanic. Details are shown for clusters with *p* <0.05. Clusters *p*-values was calculated using Kolmogorov–Smirnov test. FDR-adjusted *q* =0.2, with (1) FDR-adjusted *p* <0.2; (0) FDR-adjusted *p* ≥0.2.

**Table S3.** Untargeted semi-quantification data table for comparisons of obese with NAFL (NAFL) vs. lean healthy control (HC) in Hispanic (HIS) and White Caucasian (CAU). Details are shown for cluster members, correlations and variable importance into the projection (VIP) scores for cluster components; ANCOVA *p*-values for individual metabolites; the least square means (standard error range); and fold change (FC). Values in the table are rounded to 3 significant figures. Least square means and *p*-values were generated by age and sex adjustment.

**Table S4.** ChemRich enrichment statistics for plasma comparisons of healthy control Hispanic vs. White Caucasian. Details are shown for clusters with *p* <0.05. Clusters *p*-values was calculated using Kolmogorov–Smirnov test. FDR-adjusted *q* =0.2, with (1) FDR-adjusted *p* <0.2; (0) FDR-adjusted *p* ≥0.2.

**Table S5.** Untargeted semi-quantification data table for comparisons of healthy control subjects between ethnicities. Comparison of plasma untargeted metabolomic profile of healthy control Hispanic (HC-HIS) vs. healthy control White Caucasian (HC-CAU). Details are shown for t-test *p*-values; the least square means (standard error range); and fold change (FC). Values in the table are rounded to 3 significant figures. Least square means and *p*-values were generated by age and sex adjustment.

**Table S6.** ChemRich enrichment statistics for liver comparisons of NASH vs. 0-NASH in Hispanic. Details are shown for clusters with *p* <0.05. Clusters *p*-values was calculated using Kolmogorov–Smirnov test. FDR-adjusted *q* =0.2, with (1) FDR-adjusted *p* <0.2; (0) FDR-adjusted *p* ≥0.2.

**Table S7.** ChemRich enrichment statistics for liver comparisons of NASH vs. 0-NASH in White Caucasians. Details are shown for clusters with *p* <0.05. Clusters *p*-values are calculated using Kolmogorov–Smirnov test. FDR-adjusted *q* =0.2, with (1) FDR-adjusted *p* <0.2; (0) FDR-adjusted *p* ≥0.2.

**Table S8.** Untargeted semi-quantification data table for comparisons of liver metabolomic profiles in NASH vs. 0-NASH in both ethnicities. Details are shown for liver cluster members, correlations, *p*-values; the least square means (standard error range); fold change (FC); and ANCOVA (*t*-test) *p*-values for individual metabolites. Values in the table are rounded to 3 significant figures. Least square means and *p*-values were generated by BMI, age and race adjustment.

**Table S9.** Results from pathway analysis performed on liver metabolites of interaction with ethnicity in NASH. Metabolites with (*p*-value <0.05) on (ethnicity x NASH) from ANCOVA were mapped against pathway-associated metabolites sets from Kyoto Encyclopedia of Genes and Genomes (KEGG). Fisher’s Exact test was used for over-representation analysis, and the relative betweenness centrality was used for topology analysis. All *p*-values from enrichment analysis are adjusted for multiple testing using Benjamini-Hochberg false discovery rate (FDR) adjustment with *q* =0.2. “Total” represents the total number of compounds in the pathway; “Hits” represents the matched number of compounds detected from our data; “Impact” represents the pathway impact value calculated from pathway topology analysis.

**Table S10.** ChemRich enrichment statistics for plasma comparisons of NASH vs. 0-NASH in Hispanics. Details are shown for clusters with *p* <0.05. Clusters *p*-values was calculated using Kolmogorov–Smirnov test. FDR-adjusted *q* =0.2, with (1) FDR-adjusted *p* <0.2; (0) FDR-adjusted *p* ≥0.2.

**Table S11.** ChemRich enrichment statistics for plasma comparisons of NASH vs. 0-NASH in White Caucasian. Details are shown for clusters with *p* <0.05. Clusters *p*-values was calculated using Kolmogorov–Smirnov test. FDR-adjusted *q* =0.2, with (1) FDR-adjusted *p* <0.2; (0) FDR-adjusted *p* ≥0.2.

**Table S12.** Untargeted semi-quantification data table for comparisons for plasma metabolomic profiles in NASH vs. 0-NASH in both ethnicities. Details are shown for plasma cluster members, correlations, *p*-values; the least square means (standard error range); fold change (FC); and ANCOVA (*t*-test) *p*-values for individual metabolites. Values in the table are rounded to 3 significant figures. Least square means and *p*-values were generated by BMI, age and race adjustment.

**Table S13.** Spearman’s rank correlation analysis performed on liver metabolites and histological features in NASH.

**Table S14.** Spearman’s rank correlation analysis performed on plasma metabolites and histological features in NASH.

**Table S15.** The minimum sample size required to detect differences between group means with 80% power and 95% confidence level was calculated. Calculation is based on selected metabolites with ethnicity interaction (p <0.05) for (a) Lean healthy comparison, (b) NASH comparison.

## Notes

### Competing Interest Statement

The authors have declared no competing interest.

### Author Declarations

Institutional Review Board at the University of California, Davis (protocol # 856052).

