## Supplementary figures and images for "Ethnicity-specific alterations of plasma and hepatic lipidomic profiles are related to high NAFLD rate and severity in Hispanic Americans, a pilot study"

### Supplemental Figure 1.tiff

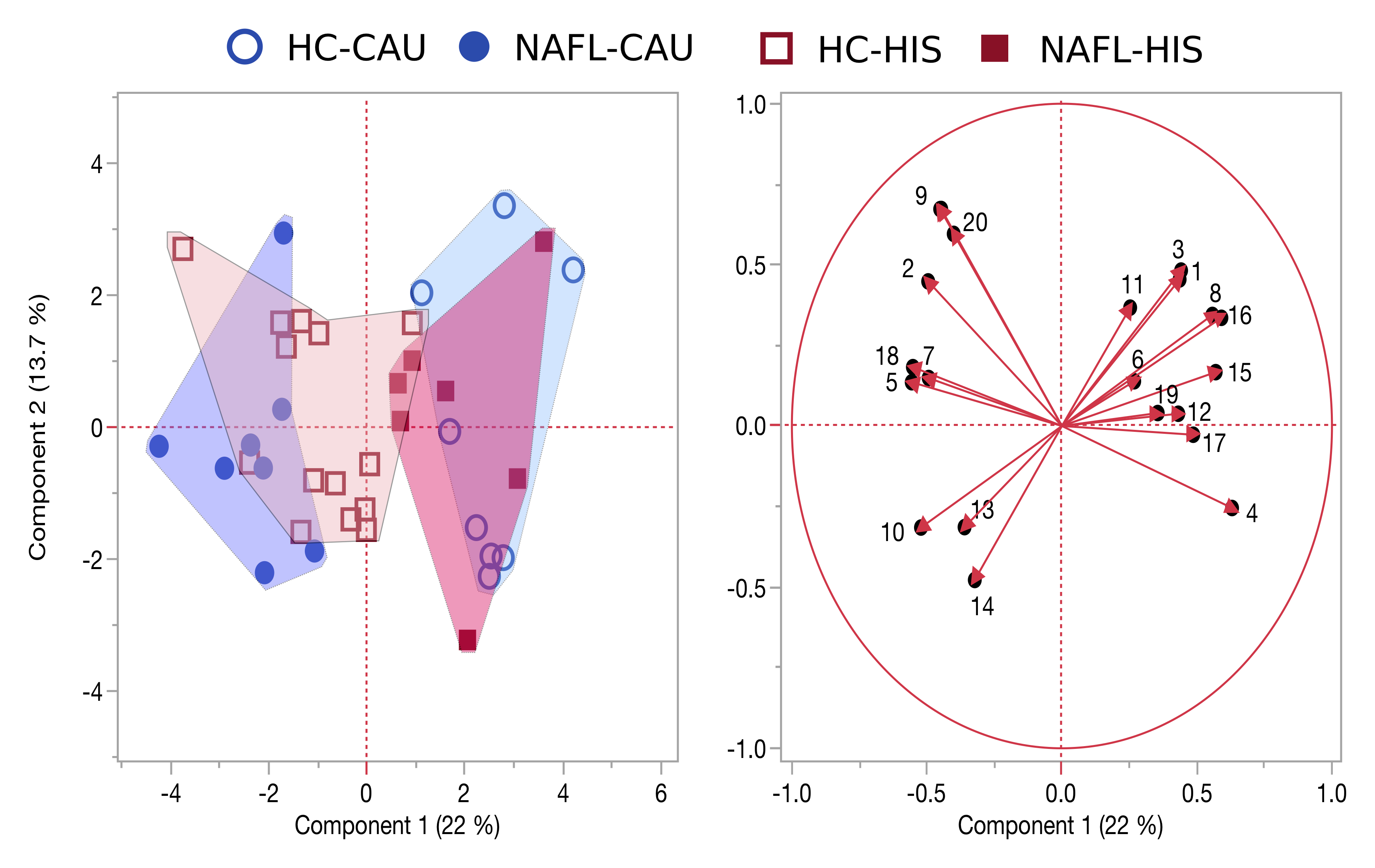
